# An interpretable, formally verified point-of-care ultrasound risk equation for difficult videolaryngoscopy: development and internal validation

**DOI:** 10.64898/2026.08.28.26361621

**Authors:** Ricardo Oyarzún-Silva, Pablo Hernández-Hernández, Miguel Ángel Fernández-Vaquero, Nekari De Luis-Cabezón

## Abstract

**Background:** Videolaryngoscopy still requires adjuncts or hyperangulated rescue in a clinically important minority, and bedside screening discriminates modestly. Point-of-care ultrasound (POCUS) of the anterior airway is a promising alternative, but existing prediction models are opaque or assume a pre-specified functional form. We developed and internally validated a parsimonious, fully disclosed POCUS risk equation whose form is recovered from data and whose structural properties are machine-checked by formal proof — to our knowledge the first formally verified clinical risk predictor — following TRIPOD+AI 2024.

**Methods:** In a prospective single-centre, single-operator cohort of 259 adults undergoing elective videolaryngoscopy (no-Easy airway 68/259, 26.3%), Sequentially Thresholded Least Squares with bootstrap stability selection (*B* = 300) screened a 71-term library of nine POCUS features and retained a seven-term logistic equation; a two-term bootstrap-stable model was pre-specified as robustness analysis. Internal validation used 5 × 10 repeated cross-validation plus temporal and device hold-outs, with pre-specified overfitting and optimism assessments. Five behavioural properties of the deployed equation were machine-checked in Lean 4.

**Results:** Two interactions met the |*c*|*/σ_c_ >* 2 stability criterion: skin-to-epiglottis × skin-to-hyoid-bone distance and tongue volume × sagittal tongue area. The seven-term equation reached a 5 × 10 cross-validated C-statistic of 0.966 (optimism-corrected 0.968) and held across temporal and device hold-outs (0.94–0.97). Calibration-in-the-large matched prevalence, with cross-validated slope 0.90 attenuating to 0.625 out-of-time; standard recalibration restored 0.92 without loss of discrimination. The pre-specified two-term robustness model reproduced this performance (C-statistic 0.964–0.968; events-per-parameter 34; shrinkage 0.99), confirming the result is not an artefact of the screening stage. Net benefit over a clinical baseline was positive across 10–50% thresholds. All five Lean 4 theorems compiled without `sorry`.

**Conclusions:** A sparse, formally verified POCUS equation predicts difficult videolaryngoscopy with high internally validated discrimination and quantified, modest overfitting. Because the equation was developed in a single-operator cohort and its inputs are operator-dependent, external validation requires prior harmonisation of the measurement protocol and operator credentialing.

## 1 Introduction

Reliable preoperative prediction of difficult videolaryngoscopy could support rational airway planning, yet most established bedside tools were derived for direct laryngoscopy, offer limited videolaryngoscopy-specific guidance, and discriminate only modestly. Developing a better predictor confronts the methodological problems that define prognostic-model research: whether the functional form is pre-specified or recovered from data, whether the fitted model is identifiable and transparently reported, whether structure selection and optimism are controlled when events per variable are low, and whether predicted risks remain calibrated out of sample.[1–3] A second, increasingly salient concern is that high-performing predictors are often opaque, and this opacity creates a trust gap that is widely held to impede safe clinical deployment.[4] Reporting standards such as TRIPOD+AI address the *transparency of reporting*—how a model is developed, validated and described—rather than the transparency of its internal mechanism;[3] an inherently interpretable model and a black box can in principle be reported with equal rigour under the same checklist, so transparent reporting and transparent mechanism remain distinct concerns.

A common response is post-hoc explainability. Attribution methods such as SHAP[5] and LIME[6] produce local approximations of a fixed black box, but they explain rather than constrain: the explanation is an estimate that can be unstable across perturbations and seeds, and it certifies no property of the underlying function. For high-stakes clinical decisions, Rudin argues that one should prefer models that are inherently interpretable rather than black boxes rationalised after the fact.[4]

We submit that the prevailing framing of the problem—“opaque vs. interpretable”—is too narrow. Three families of predictive models differ in the *methodological rigor* with which they constrain the claims a fitted model is licensed to make. Regularised logistic regression estimates coefficients under a functional form that is fixed before the data are seen, so the hypothesis it tests is restricted to a pre-specified linear-additive structure. Gradient-boosted trees fit a high-capacity ensemble that lacks an identifiable closed form and admits no audit of its behaviour on inputs outside the training distribution. Sparse symbolic regression coupled with formal verification recovers a fully identifiable closed-form expression from a structured candidate space, exposes the selection itself to bootstrap-stability checks, and certifies properties of the fitted function over its entire feasibility region. In high-stakes clinical decisions the question is therefore not only which model discriminates best, but which model carries the strongest auditable warrant for the claims it makes about the prediction problem.

Sparse symbolic regression offers interpretability by construction. Methods in the family of sparse identification of nonlinear dynamics[7] and its ensemble extension E-SINDy,[8] together with related universal differential equation formulations,[9] recover compact closed-form expressions from a candidate library. Methodologically, the form of the model is *recovered* from a structured space of candidate terms rather than fixed beforehand: which interactions and which transformations belong to the predictor becomes a testable outcome of inference, with selection uncertainty quantifiable by resampling-based stability [10], rather than a presupposition that is never exposed to data. Interpretability, however, is not correctness: a sparse formula remains free to violate domain or monotonicity constraints at the edges of the operating range—for example, to leave the probability simplex or to reverse a clinically established direction of effect—precisely the monotonicity and output-domain properties we later certify.

Interactive theorem provers close this remaining gap. Lean 4 with the Mathlib library[11, 12] permits machine-checked proof that a function satisfies stated mathematical properties for all admissible inputs. Such proofs certify *behavioural* properties of the predictor as a mathematical object—its output domain, its monotonicities and the sign of specified effects—over the full feasibility region of inputs, and not its empirical performance, calibration, clinical safety or generalisation, which remain matters for data. The distinction is methodologically consequential: behavioural properties established by proof apply by construction to every admissible input, whereas empirical performance metrics estimate sample averages [1, 2]. To our knowledge, and on a targeted search of the clinical-AI and formal-methods literature, no prior clinical risk predictor has had such behavioural properties machine-checked.

We report what is, to our knowledge, the first machine-checked behavioural-property verification of a clinical risk predictor, in a single-centre, single-operator point-of-care ultrasound (POCUS) case study. The cohort comprises *n* = 259 patients from a single centre (Hospital Universitario de Navarra, Madrid), with all ultrasound measurements acquired by one operator; the predicted outcome is a binary airway-management difficulty label (*Easy* versus *no-Easy*, prevalence 26.3%). The predictor is a sparse closed-form formula built from quantitative airway-ultrasound measurements (nine POCUS indices). This work extends a previously reported multimodal model developed on an earlier freeze of the same single-operator cohort [13], which addressed a three-class videolaryngoscopy-strategy outcome with conventional tree-ensemble, support-vector and multinomial-regression algorithms (independent test-set macro-averaged AUC 0.95); the present study is methodologically distinct in four respects: it re-casts the task as a binary *Easy*/*no-Easy* prediction, incorporates additional ultrasound-derived parameters, replaces black-box algorithms with a sparse symbolic regression that yields a disclosed closed-form equation, and adds formal verification together with a TRIPOD+AI-conformant internal-validation, overfitting and calibration assessment. This design entails the limitations that TRIPOD+AI foregrounds—a single centre, dependence on a sole sonographer, and the absence of external or geographic validation—which we state plainly here and revisit in the Discussion. Our contributions are threefold: (i) a sparse-regression pipeline coupling sequentially thresholded least-squares with bootstrap stability selection to yield a parsimonious, interpretable formula that constitutes a falsifiable mechanistic conjecture rather than only a fitted model; (ii) five Lean 4 theorems that certify the predictor’s monotonicity, output-domain (0, 1) and sign-of-effect properties (including the antitone dependence of the scoring function on thyromental distance, a property of the fitted function not a clinical claim) for all admissible inputs; and (iii) a TRIPOD+AI-conformant internal-validation, overfitting and calibration assessment showing that the interpretable model matches strong black-box comparators within this cohort while additionally yielding machine-checkable structural guarantees on the deployed equation that those comparators do not provide.

## 2 Methods

### 2.1 Cohort

Consecutive adult patients undergoing elective surgery with general anaesthesia and orotracheal intubation at the Hospital Universitario de Navarra (Madrid, Spain) were prospectively enrolled from January 2023 in an ongoing single-centre registry (ClinicalTrials.gov NCT06925009); the present analysis is based on a data freeze of April 2026. Inclusion required age ≥ 18 years, planned videolaryngoscopy-assisted intubation, and informed consent. Exclusion criteria were emergency surgery, awake intubation, cervical immobilisation precluding ultrasound assessment, and known major airway pathology distorting baseline sonoanatomy. Of 260 enrolled patients, 1 was excluded for an incomplete ultrasound dataset, leaving *n* = 259 with full predictor and outcome data for analysis; the single excluded patient was not imputed, and no predictor values were imputed for the analysed cohort, so all analyses use complete cases. This cohort is an updated, extended extraction of the same single-operator dataset reported in the parent Airway Coach study [13]: it enrols additional consecutively recruited patients acquired after that report’s data freeze and incorporates ultrasound-derived parameters (tongue volume and tongue width) not modelled there; minor differences in patient counts and grade frequencies relative to the parent reflect the later freeze and are expected for an ongoing prospective registry.

All point-of-care ultrasound (POCUS) acquisitions and clinical airway examinations were performed by a single anaesthesiologist (M.A.F.-V.) immediately prior to induction. This design eliminates between-operator variability but, by the same token, leaves the inter-operator reproducibility of the nine POCUS measurements unvalidated; we treat this as a first-order generalisability threat and return to it in the Discussion. Because the same operator acquired the ultrasound measurements while the difficulty grade was not formally blinded to those measurements, a degree of incorporation bias cannot be excluded; this is mitigated by anchoring the outcome to objective procedural events (the need for intubation adjuncts or escalation to a hyperangulated device) under the Video Classification of Intubation framework rather than to a subjective impression, and we flag the residual risk explicitly. Intubations were performed with either the McGrath MAC (*n* = 132) or the C-MAC D-Blade (*n* = 127) videolaryngoscope, allocated according to clinical workflow rather than randomisation, which enabled the device hold-out analysis described below. The study was approved by the Research Ethics Committee of the University of Navarra and registered at ClinicalTrials.gov (NCT06925009), and was conducted in accordance with the Declaration of Helsinki; full committee details, protocol number and approval date are given in the Ethics approval and consent to participate declaration.

The primary outcome was the operator-rated Airway Coach grade (0 = uneventful first-pass intubation; 1 = first pass with adjuncts or repositioning; 2 = failed first pass requiring additional manoeuvres). For the binary classification task this grade was dichotomised as *Easy* (grade 0) versus *no-Easy* (grades 1–2). The cohort comprised *n* = 191 Easy (73.7%) and *n* = 68 no-Easy events (26.3%), of which *n* = 51 were grade 1 and *n* = 17 grade 2.

### 2.2 Predictor variables

Nine sonographic features were prospectively acquired with a linear high-frequency probe in standardised planes: distance from skin to epiglottis (dse); distance from skin to hyoid bone (dshb); tongue thickness in the midsagittal plane (tt); tongue volume, computed as the product of sagittal tongue area and tongue width (tvol); sagittal tongue area, the cross-sectional area of the tongue in the sagittal plane (star); thyromental distance measured sonographically (tmd); sternomental distance (smd); tongue width in the transverse plane (tw); and the hyomental distance ratio in neutral versus extended neck position (hmdr). The clinical rationale follows established sonoanatomical work on the difficult airway: anterior-neck depth measurements (dse, dshb) capture the soft-tissue column between skin and airway that the blade must displace, lingual-mass measurements (tt, tvol, star, tw) capture the tongue volume occupying the oral cavity, geometric ratios (hmdr) capture airway compressibility under neck extension, and skeletal distances (tmd, smd) capture the mandibular space available for tongue displacement. All measurements were recorded as continuous variables in their native units.

For benchmarking, a clinical baseline feature set was assembled from established bedside predictors: age, body mass index (BMI), modified Mallampati score (MMS), upper-lip bite test (ULBT), neck circumference (NC), bedside thyromental distance (TMD_clin_), inter-incisor distance (IID), and biological sex.

### 2.3 Sparse symbolic regression via STLSQ

We treat the discovery of a parsimonious predictor as a sparse symbolic regression problem in the spirit of SINDy [7] and its ensemble extension E-SINDy [8]. Let **x***_i_* ∈ R^9^ denote the POCUS feature vector for patient *i* and *y_i_* ∈ {0, 1} the binarised outcome. We construct a feature library Θ(**x**) comprising the nine original features, their squares, all pairwise products *x_j_x_k_*for *j < k*, eight pre-specified anatomical ratios, and natural logarithms, giving a candidate dictionary of 71 terms (9 linear + 9 quadratic + 36 pairwise products + 8 ratios + 9 logarithms). To control the pathologies of ratio and logarithmic transforms, raw measurements are first winsorised at the 1st/99th percentiles to bound outliers; ratios and logarithms are formed on the raw (strictly positive) measurement scale, with denominators floored away from zero, *before* the resulting library columns are standardised to zero mean and unit variance. Ratios are therefore ratios of measurements rather than ratios of *z*-scores, and the standardisation is applied once, downstream of all non-linear feature construction. The candidate model is

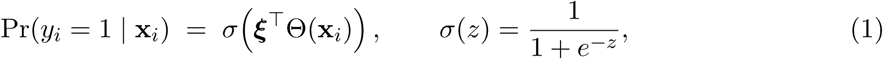

where ***ξ*** is the coefficient vector we seek to recover, ideally sparse. Sparsity is induced by Sequentially Thresholded Least Squares (STLSQ) adapted to the logistic loss: at each iteration we fit an *L*_1_-penalised binomial deviance, hard-threshold coefficients with |*ξ_j_*| *< τ* to zero, and refit the surviving support without penalty, iterating until the active set is stable. The high-level procedure (input: library Θ, outcome **y**, sparsity threshold *τ*, penalty *λ*; repeat fit–threshold–refit until the support is unchanged) is given as runnable pseudocode in the Supplementary Material, sufficient for independent re-implementation; exact values of *λ* and *τ* are available under the research collaboration agreement, while random seeds are included in the released code described below.

We emphasise the events-per-variable (EPV) tension this creates. The 71-term candidate library is screened against only 68 no-Easy events — an events-per-candidate ratio of approximately 0.96 during selection, far below conventional EPV targets and below the minimum sample size that the Riley et al. criteria would prescribe for a model of this apparent dimensionality [1, 3]. Selection-induced optimism is therefore a material risk that sparsity alone does not remove. To mitigate it we apply bootstrap stability selection in the E-SINDy style [8]: *B* = 300 stratified bootstrap resamples are drawn, the *entire* STLSQ pipeline (including the selection step, not merely coefficient estimation) is refitted on each resample, and only library terms whose bootstrap coefficient-to-standard-deviation ratio satisfies |*ξ̅j*|*/*SD(*ξ_j_*) *>* 2 are treated as load-bearing. Seven terms were retained for the apparent model; two met the stability criterion. The most stable was the anterior-neck-depth interaction dse×dshb (skin-to-epiglottis × skin-to-hyoid-bone; *c̅* = +1.68 ± 0.54, |*c̅*|*/*SD = 3.1), followed by the lingual-mass interaction tvol×star (tongue volume × sagittal tongue area; *c̅* = +1.57 ± 0.71, |*c̅*|*/*SD = 2.2). The first interaction is composed of two anterior-neck depth measurements and is interpreted as such; it does not involve tongue thickness and is not a lingual-mass construct. Only the second interaction is a lingual-mass term. The remaining five terms (star/tt, dse×tvol, tmd×hmdr, dse^2^, log tmd) did not reach the threshold and are reported as transparent secondary structure rather than as established predictors. The seven-term model is the deployed equation (Section 3.6); we additionally pre-specified a parsimony analysis restricting the model to the two bootstrap-stable interactions as a robustness check, and quantified fitting-stage overfitting by events-per-estimated-parameter, the van Houwelingen heuristic shrinkage factor, the Riley (2019) minimum-sample-size criteria, and Harrell–Steyerberg bootstrap optimism in both the C-statistic and the calibration slope (Section 3.6) [1, 2].

### 2.4 Formal verification in Lean 4

The retained symbolic predictor was transcribed into Lean 4 [11, 12] as a total function `predict` : `Features` → R, with `Features` a structure of nine real fields and positivity assumed as an explicit hypothesis wherever a theorem requires it. Five theorems were *stated* and machine-checked; the complete proof of **P1** (sigmoid monotonicity) is additionally reproduced in the Supplementary Material as a worked example, and the complete proof project for all five theorems is released in the public code repository (https://doi.org/10.5281/zenodo.22143665). **P1** (sigmoid monotonicity): `predict` is strictly increasing in the linear score. **P2** (domain): `predict` is well-defined and lies in (0, 1) for all strictly positive feature inputs, with no division-by-zero or log-of-non-positive pathologies. **P3** (tmd direction-of-effect): with the remaining features held fixed and the hyomental-distance ratio positive, `predict` is strictly antitone in tmd on its positive domain; tmd enters only through the tmd×hmdr and log tmd terms, both carrying negative coefficients. **P4–P5**: the contribution of each bootstrap-stable interaction (dse×dshb, tvol×star) to the score is strictly increasing in the interaction value, in the direction of its positive coefficient. The whole project compiles (lake build, Lean 4.30.0 with Mathlib) with zero sorry placeholders.

We are deliberately precise about what this establishes and what it does not. The theorems certify mathematical properties—monotonicity, codomain, sign-consistency—of a *fixed-coefficient* function whose bootstrap-estimated signs are hard-coded as literals. They do *not* verify that those signs are clinically correct, that the model is well specified, or any physiological fact. In particular P3 certifies that the deployed function—with its data-estimated negative coefficients on the two tmd terms—is monotone-decreasing in tmd; because those signs were fitted rather than imposed, the theorem largely follows from them and is reported as a self-consistency guarantee on the deployed artefact (it cannot be silently broken by refactoring, retraining on the same library, or numerical re-implementation), not as empirical evidence that tmd is clinically protective.

### 2.5 Calibration

Because sparse fitting used class-balanced weights to stabilise selection, the final coefficient vector was refitted *unweighted*on each training partition so that the mean predicted risk matches cohort prevalence by construction (a prevalence-true intercept) [1, 2]. Our primary calibration evidence is calibration-in-the-large expressed as mean predicted probability versus observed prevalence, accompanied by the calibration slope, the Brier score, and a reliability diagram. We do not headline the Cox calibration-in-the-large intercept as a single signed number: in highly discriminative models it is inflated by a Jensen-type gap and is an unreliable miscalibration signal; the Cox value is relegated to Supplementary material with that caveat. On the pooled 5 × 10 cross-validation, mean predicted risk was 26.2% against an observed 26.3%, with slope 0.904 and Brier 0.033. On the 80/20 test set, mean predicted risk was 26.5% against observed 26.9% and Brier was 0.028, but the calibration slope was 0.714. A slope of 0.71 means the linear predictor is materially over-extreme out of sample—roughly 30% shrinkage—and is a genuine miscalibration signal on a held-out set rather than “near unity”; we report it explicitly and treat post-deployment shrinkage recalibration as required. Calibration-in-the-large and Brier are thus good (mean risk tracks prevalence; Brier well below the ∼0.19 null at 26% prevalence), while the out-of-sample slope indicates residual over-confidence that future recalibration must address.

### 2.6 Validation pipeline

We evaluated the sparse predictor against three benchmarks—a clinical-baseline logistic regression, a dense nine-feature ultrasound logistic regression, and an XGBoost model on the same library—under: (i) 5 × 10 repeated stratified cross-validation with per-fold refitting of every pipeline stage, including stability selection; (ii) a stratified 80/20 split (test *n* = 52, 14 positive, 26.9% observed); (iii) a temporal hold-out fitting on the first enrolment half and testing on the second (*n* = 129, leaving a *n* = 130 training half; patients were partitioned by order of enrolment, captured sequentially); and (iv) device hold-outs (McGrath→C-MAC, *n* = 127; C-MAC→McGrath, *n* = 132). We stress that all four partitions derive from the *same* single-centre, single-operator cohort: the temporal and device hold-outs probe out-of-sample transfer, not external, multi-operator, or multi-centre validity, and are not described as external validation. Optimism was estimated by the Harrell–Steyerberg bootstrap (*B* = 200), in which the *full* pipeline—feature construction *and* STLSQ selection—was re-executed inside every resample, so the reported optimism reflects selection as well as estimation. For the sparse model the apparent AUC was 0.975 and the estimated optimism 0.007, giving an optimism-corrected AUC of 0.968.

Discrimination was reported as AUC with bootstrap 95% confidence intervals, plus sensitivity and specificity at the Youden-optimal threshold; we flag that with only 68 events and test folds as small as *n* = 52/14 positives the upper CI tail is unstable and several intervals reach 1.000, so the near-perfect point estimates must be read against the low EPV and large candidate library rather than as evidence of separability. Calibration (mean-predicted-vs-prevalence, slope, Brier, reliability diagram) was computed on held-out predictions and is additionally reported for the temporal and device hold-outs, not only for pooled CV and the 80/20 split, to support any transfer claim. Reclassification against the clinical baseline used the continuous and categorical Net Reclassification Index and the Integrated Discrimination Improvement, each reported with its event and non-event components and with the standard caveats on categorical NRI at the arbitrary 0.30 threshold; the IDI is defined as the difference in discrimination slopes and, given its unusually large magnitude here (sparse IDI +0.533, with categorical NRI +0.444 on the 80/20 test), was independently recomputed to exclude a scaling error and is interpreted cautiously. Decision-curve analysis was computed on held-out (not apparent) predictions, with tabulated net-benefit values confirming positive net benefit over the ∼10–50% threshold range relative to treat-all, treat-none and the clinical baseline. Reporting follows the TRIPOD+AI 2024 statement [3]; a completed checklist, the intended use and target population, the model-lock date and version, and prespecified fairness analyses by sex and BMI are provided as Supplementary material.

### 2.7 Software and data availability

The pipeline was implemented in Python 3.11.9 (numpy 2.4.4, pandas 2.3.3, scipy 1.17.1, scikit-learn 1.8.0, statsmodels 0.14.6, xgboost 3.1.3, matplotlib 3.10.8), and the formal verification in Lean 4 (v4.30.0) with Mathlib. The complete Lean 4 proof project (all five theorems, no sorry placeholders) and the analysis and validation scripts are released in a public repository at submission, allowing independent re-verification of every reported guarantee and every reported metric. The deployable equation and its standardisation constants are disclosed in full in the manuscript. Only the exact hyperparameters of the STLSQ screening stage (*λ*, *τ*) remain available under a research collaboration agreement while a patent application covering the screening methodology is pending; these values affect model discovery only: the repository includes `verify_equation_public.py`, which reconstructs the seven disclosed terms, refits the deployed equation and checks it against both the published coefficients and the Lean constants without using them, and the Lean proof project re-verifies every formal guarantee from the repository alone, requiring neither the hyperparameters nor the data.

## 3 Results

### 3.1 Cohort characteristics

From the registry’s inception in January 2023 to the April 2026 data freeze, 260 consecutive adult patients undergoing elective surgery requiring tracheal intubation were enrolled at our single centre; one was excluded for an incomplete ultrasound dataset, leaving 259 analysed. Anatomical, clinical, and ultrasound measurements were performed by a single operator (MAFV) using a standardised protocol. By construction this design removes inter-operator variability from the present cohort; we note, however, that it is double-edged, because the performance reported here is conditional on one expert acquirer and need not transfer unchanged to other operators or centres (Discussion). Intubation was performed with the McGrath videolaryngoscope in 132 patients (51.0%) and with the C-MAC videolaryngoscope in 127 patients (49.0%). The operator-rated Airway Coach grade was G0 (Easy) in 191 patients (73.7%), G1 in 51 (19.7%) and G2 in 17 (6.6%), yielding a composite “no-Easy” airway outcome (grade ≥ 1) in 68 patients (26.3%).

Table 1 summarises demographic, clinical, ultrasound and procedural data stratified by Airway Coach difficulty grade. Patients with higher grades were older (mean age 56.5 versus 63.4 versus 66.1 years for G0/G1/G2), had higher BMI (25.2 versus 27.7 versus 28.2 kg/m^2^), and a male predominance increased monotonically with grade (47.1% versus 74.5% versus 82.4%). Classical clinical predictors followed the expected gradient: modified Mallampati III-IV prevalence rose from 6.3% in G0 to 35.3% in G2, and upper-lip bite test class III from 0.0% to 17.6%. Ultrasound markers showed clear stratification: skin-to-hyoid distance (DSHB), skin-to-epiglottis distance (DSE), tongue volume and sagittal tongue area (STAR) all increased with grade. Predictor and outcome completeness was effectively total in the analytical cohort (one of 260 enrolled patients excluded for an incomplete ultrasound dataset, leaving 259 analysed; no imputation required). The cohort flow from enrolment to the analysed dataset and the grade distribution are summarised in Figure 1.

**Table 1:** Baseline characteristics of the Navarra cohort (*n* = 259) stratified by Airway Coach difficulty grade. Values are mean (SD) or median [IQR] according to the within-stratum normality of each continuous variable (so the summary statistic may differ across grade columns for the same variable), and *n* (%) for categorical variables. G0 = Easy, G1 = intermediate, G2 = most difficult.

| Variable | G0 ($n = 191$) | G1 ($n = 51$) | G2 ($n = 17$) |
| --- | --- | --- | --- |
| *Demographics* |  |  |  |
| Age (years) | 56.49 (15.92) | 63.37 (11.39) | 66.12 (10.72) |
| BMI ($\text{kg}/\text{m}^2$) | 25.2 [22.8–27.5] | 27.67 (3.44) | 28.19 (3.17) |
| Sex – Female | 101 (52.9%) | 13 (25.5%) | 3 (17.6%) |
| Sex – Male | 90 (47.1%) | 38 (74.5%) | 14 (82.4%) |
| ASA I | 36 (18.8%) | 3 (5.9%) | 1 (5.9%) |
| ASA II | 141 (73.8%) | 43 (84.3%) | 12 (70.6%) |
| ASA III | 14 (7.3%) | 5 (9.8%) | 4 (23.5%) |
| *Clinical predictors* |  |  |  |
| TMD (cm) | 7.35 (0.59) | 7.18 (0.60) | 7.09 (0.69) |
| SMD (cm) | 13.6 [13.1–14.1] | 13.36 (0.94) | 13.49 (1.03) |
| IID (cm) | 4.09 (0.32) | 3.91 (0.39) | 3.71 (0.39) |
| NC (cm) | 38.07 (2.66) | 41.24 (3.31) | 42.59 (3.97) |
| MMS III–IV (difficult) | 12 (6.3%) | 13 (25.5%) | 6 (35.3%) |
| ULBT III (difficult) | 0 (0.0%) | 4 (7.8%) | 3 (17.6%) |
| *POCUS measurements* |  |  |  |
| DSHB (cm) | 0.97 (0.16) | 1.32 (0.22) | 1.35 (0.19) |
| DSE (cm) | 1.92 (0.30) | 2.5 [2.5–2.6] | 2.69 (0.22) |
| HMDn (cm) | 4.89 (0.51) | 5.12 (0.56) | 5.16 (0.65) |
| HMDe (cm) | 5.59 (0.55) | 5.66 (0.66) | 5.58 (0.57) |
| HMD ratio | 1.14 (0.06) | 1.10 (0.06) | 1.1 [1.0–1.1] |
| STAR ($\text{cm}^2$) | 19.54 (2.45) | 25.88 (2.70) | 26.8 [25.5–28.0] |
| TT (cm) | 5.42 (0.62) | 5.99 (0.74) | 6.18 (0.83) |
| TW (cm) | 4.38 (0.57) | 5.08 (0.64) | 5.42 (0.43) |
| Tongue volume ($\text{cm}^3$) | 85.40 (19.81) | 132.34 (25.72) | 148.68 (32.19) |
| *Videolaryngoscope* |  |  |  |
| VL – McGrath | 98 (51.3%) | 27 (52.9%) | 7 (41.2%) |
| VL – C-MAC | 93 (48.7%) | 24 (47.1%) | 10 (58.8%) |

**Figure 1:**
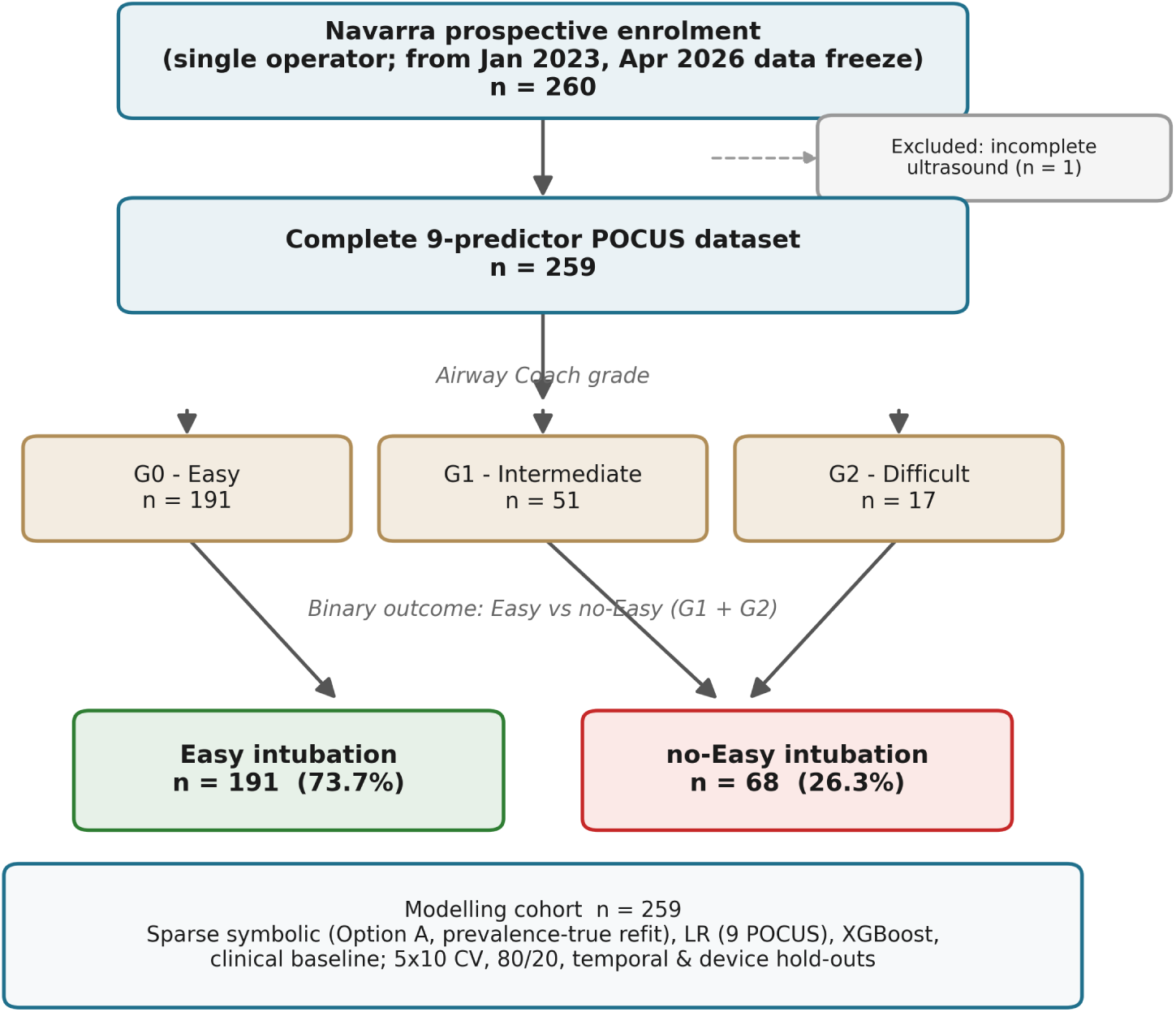
Flow of the Navarra single-centre cohort from prospective enrolment (n=260; single operator; from January 2023, April 2026 data freeze) through exclusion of one patient with incomplete ultrasound to the complete 9-predictor POCUS dataset (n=259), the Airway Coach grade distribution (G0=191, G1=51, G2=17), and the binary modelling outcome of Easy (n=191, 73.7%) versus no-Easy intubation (n=68, 26.3%).

### 3.2 Sparse symbolic predictor: structure and stability

Sequentially Thresholded Least Squares (STLSQ) applied to the full feature library (the nine ultrasound base features plus their squares, all pairwise products, eight pre-specified ratios and logarithms; 71 candidate terms) retained seven terms in the final symbolic model. The events-per-candidate ratio during selection is the conservative figure: 68 events over 71 candidate terms equals approximately 0.96, well below the conventional Riley-Steyerberg targets and the precise regime in which selection cannot be governed by apparent fit [1, 2]. Bootstrap stability selection (*B* = 300, |*c*|*/σ_c_ >* 2) is therefore the constraint that controls the chosen model; the resulting seven-term predictor and the nine-predictor benchmark logistic regression then correspond to approximately 7.5–9.7 events per estimated parameter at the post-selection *fitting* stage, at or slightly below the conventional target. Two terms reached the pre-specified stability criterion |*c*|*/σ_c_ >* 2 under bootstrap stability selection (*B* = 300): the product DSE×DSHB (bootstrap mean coefficient +1.68 ± 0.54; |*c*|*/σ_c_* = 3.13) and the product TVOL×STAR (bootstrap mean coefficient +1.57±0.71; |*c*|*/σ_c_* = 2.21). The bootstrap mean and standard deviation are computed over *B* = 300 resamples and quantify selection-level uncertainty; the corresponding full-cohort point-estimate coefficients deployed in Eq. (2) below are +1.613 and +1.458 respectively, each within one bootstrap standard deviation of its mean. The remaining five terms were retained in the apparent model but did not meet the stability threshold and are reported as a transparent secondary structure rather than load-bearing components of the predictor; the intercept itself reached |*c*|*/σ_c_* = 3.93 on the bootstrap distribution. The standardized log-odds coefficients of the seven retained terms and their bootstrap stability are shown in Figure 4.

Both stable terms admit a direct anatomical interpretation. The DSE×DSHB product couples the depth of the epiglottis with the depth of the hyoid, jointly characterising the anterior soft-tissue column between the skin and the airway: an enlargement on either axis increases the distance the laryngoscope blade must displace to expose the glottis. This is a pre-laryngeal soft-tissue construct and does not involve tongue thickness (TT). The TVOL×STAR product, by contrast, couples tongue volume with the sagittal tongue area, capturing the lingual mass that must be displaced to clear the line of sight; this term is the lingual-mass interaction proper. Both terms carry positive coefficients consistent with the expected direction of clinical risk for airway difficulty.

### 3.3 Final model specification

The deployable predictor is fully disclosed here. Let *z*(·) denote standardisation *z*(*t*) = (*t* − *µ_t_*)*/s_t_*applied to each constructed library term *t*, with the constants (*µ_t_, s_t_*) given in Table 2. The full-cohort fit (*n* = 259, Option-A standardised logistic model) has linear predictor

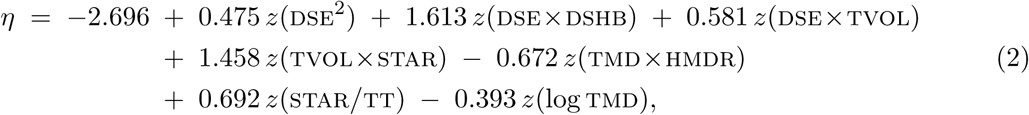

**Table 2:** Standardisation constants for the seven retained library terms in Eq. (2). Each term *t* is standardised as *z*(*t*) = (*t* <u>−</u> *<u>µ_t_</u>*)*<u>/s_t_</u>* <u>before entering the linear predi</u>ctor.

| Term | Mean ($\mu_t$) | Scale ($s_t$) |
| --- | --- | --- |
| DSE<sup>2</sup> | 4.5277 | 1.6952 |
| DSE × DSHB | 2.2951 | 0.9016 |
| DSE × TVOL | 214.975 | 105.337 |
| TVOL × STAR | 2216.455 | 1175.663 |
| TMD × HMDR | 8.2699 | 0.8574 |
| STAR/TT | 3.8260 | 0.5774 |
| log TMD | 1.9848 | 0.0813 |

and the predicted risk follows the logistic link

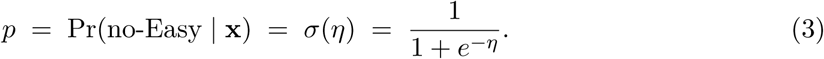

Lengths (dse, tt, tmd, and the hmdr components) are in cm, tvol in cm^3^, star in cm^2^; log denotes the natural logarithm. The full-cohort point estimates deployed in Eq. (2) (+1.613 for dse×dshb and +1.458 for tvol×star) and the bootstrap means reported in §3.2 (resp. +1.68 ± 0.54, |*c*|*/*SD = 3.13; +1.57 ± 0.71, |*c*|*/*SD = 2.21) reflect two distinct quantities computed on the same library: the deployed equation uses the unique point estimate from the single full-cohort logistic refit, while the bootstrap statistics quantify the selection-level sampling variability over *B* = 300 resamples. The two are mutually consistent: each point estimate lies within one bootstrap standard deviation of the corresponding mean, supporting the stability finding. Similarly, the intercept −2.696 in Eq. (2) is the point estimate from the full-cohort fit, whereas −3.003 ± 0.76 is the bootstrap-mean intercept reported in the supplementary stability table.

Equations (2)–(3) together with Table 2 constitute the complete deployable equation, disclosed in full so that any reader can compute a risk estimate from the nine ultrasound measurements; the complete Lean 4 proof project and the analysis scripts are released in the public code repository, and only the STLSQ screening hyperparameters (the sparsity threshold *τ* and the regularisation path *λ*) remain under a research collaboration agreement (patent pending).

### 3.4 Discrimination

Discrimination metrics are summarised in Table 3. Under 5×10 stratified cross-validation, the sparse symbolic predictor achieved per-fold mean AUC 0.966 (95% CI 0.926–1.000), with a pooled-predictions AUC of 0.963; the two summaries differ slightly because the per-fold mean averages the 50 fold-level AUCs whereas the pooled estimate computes a single AUC on all out-of-fold predictions concatenated. Both are numerically close to *ℓ*_2_-regularised logistic regression on the nine ultrasound predictors (AUC 0.967) and to gradient-boosted trees (XGBoost AUC 0.955), with overlapping confidence intervals among the three. All three ultrasound-based models exceeded a clinical baseline of eight standard bedside predictors (age, BMI, sex, modified Mallampati, ULBT, neck circumference, thyromental distance and inter-incisor distance; AUC 0.809), an absolute gain of approximately 0.15–0.16 in AUC. We emphasise that overlapping intervals do not establish statistical equivalence: no formal equivalence test or paired DeLong contrast was pre-specified on these small samples, so the three top models are best read as numerically comparable rather than as proven indistinguishable, and the apparent absence of a discrimination penalty for the interpretable formula is an observation rather than a tested claim. The upper confidence bound of 1.000 for all three ultrasound models, together with the XGBoost apparent AUC of exactly 1.000 reported below, is consistent with near-separability of the outcome on *n* = 259 with 68 events and signals that these point estimates are imprecise at the upper end.

**Table 3:** Discrimination across validation settings. Area under the ROC curve (AUC) for the sparse symbolic model (with bootstrap 95% CI where available), the nine-predictor logistic regression (LR), the XGBoost reference model, and the clinical-baseline model; the rightmost column reports sensitivity / specificity for the sparse model at the Youden-optimal threshold on each setting’s predictions. The optimism-corrected row applies the Harrell–Steyerberg bootstrap (*B* = 200) to the sparse model.

| Setting | Sparse AUC [95% CI] | LR AUC | XGB AUC | Clin AUC | Sparse Sens/Spec @ Youden |
| --- | --- | --- | --- | --- | --- |
| 5 × 10 repeated CV | 0.966 [0.926–1.000] | 0.967 | 0.955 | 0.809 | 0.91 / 0.98 |
| 80/20 test ($n = 52$) | 0.930 | 0.936 | 0.932 | 0.746 | 0.93 / 1.00 |
| Temporal 1H→2H ($n = 129$) | 0.941 | 0.943 | 0.948 | – | 0.87 / 0.99 |
| Device Mc→C-MAC ($n = 127$) | 0.947 | 0.948 | 0.949 | – | 0.91 / 0.98 |
| Device C-MAC→Mc ($n = 132$) | 0.972 | 0.978 | 0.969 | – | 0.94 / 0.97 |
| Optimism-corrected | 0.968 | – | 0.984 | – | – |

CI = bootstrap 95% confidence interval; Mc = McGrath. Sens/Spec @ Youden are reported at the threshold that maximises Youden’s $J = \text{sensitivity} + \text{specificity} - 1$ on each setting’s held-out predictions for the sparse model (CV row uses pooled out-of-fold predictions). Optimism-corrected sparse: apparent 0.975, optimism 0.007, corrected 0.968; XGBoost apparent 1.000 (in-cohort overfit reference). Dashes denote cells omitted for brevity or not applicable: the clinical-baseline AUCs for the transfer rows (0.76–0.81) are reported in the Supplementary Material, and the optimism-corrected and Youden-threshold metrics are defined only for the rows shown.

On the 80/20 stratified hold-out split (test *n* = 52, 14 events) the sparse model reached AUC 0.930 (95% CI 0.778–1.000); the benchmark logistic regression reached 0.936 and XGBoost 0.932. Robustness to distributional shift was probed by three further internal splits, all single-centre and single-operator and each reported with bootstrap 95% confidence intervals. Under the temporal split (first half of inclusion as training, second half as test, *n* = 129) the sparse model achieved AUC 0.941 (0.857–0.995), consistent with the cross-validated estimate. Under the device hold-outs the sparse model reached AUC 0.947 (0.863–0.997) trained on McGrath and tested on C-MAC (*n* = 127), and 0.972 (0.930–0.999) in the reverse direction (*n* = 132). These are internal partitions of one cohort acquired by one operator and thus evidence of stability across recruitment period and device within this setting; they are not cross-device external validation, and the wide intervals on each small hold-out preclude a strong claim of device invariance. The ROC curves with bootstrap confidence bands across the three transfer settings are shown in Figure 2.

**Figure 2:**
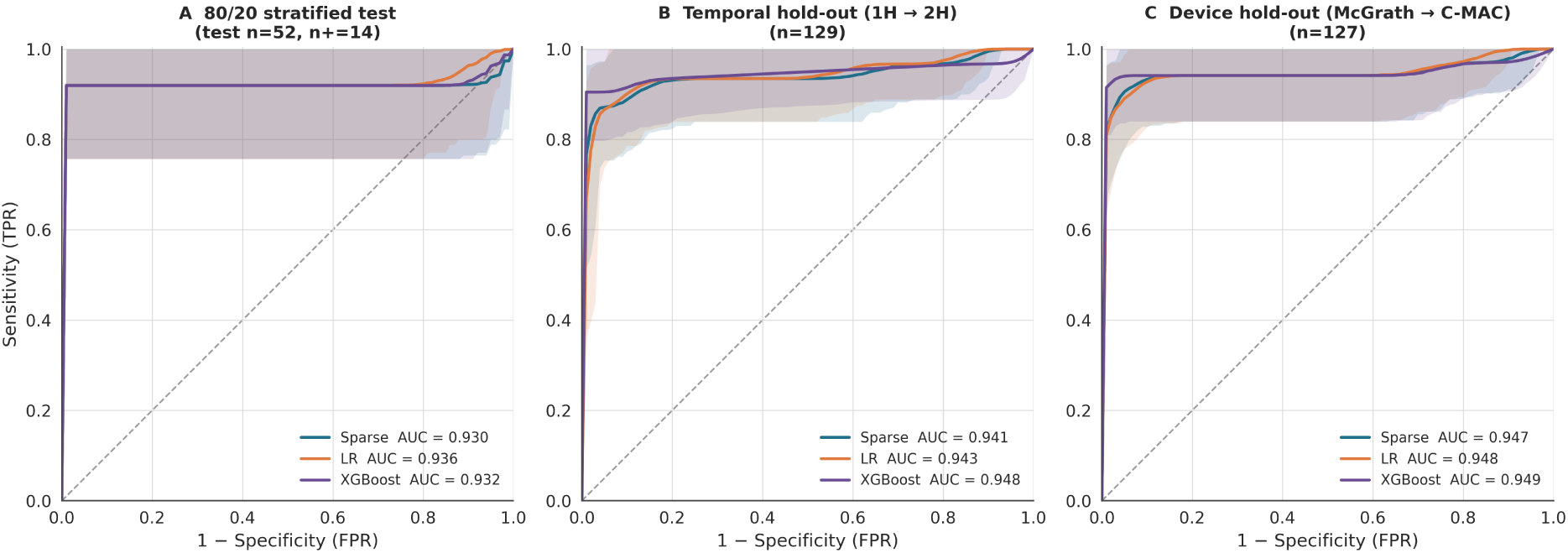
Receiver-operating-characteristic curves with bootstrap 95% confidence bands (B=300) for the sparse symbolic, logistic-regression and XGBoost models across three validation settings — 80/20 stratified test (Sparse AUC 0.930, LR 0.936, XGBoost 0.932), temporal 1H→2H hold-out (0.941/0.943/0.948) and McGrath→C-MAC device transfer (0.947/0.948/0.949) — showing near-identical, well-discriminating performance of the interpretable model relative to the black-box benchmarks.

### 3.5 Optimism correction

Harrell–Steyerberg bootstrap optimism correction (*B* = 200) yielded an apparent AUC of 0.975 and a corrected AUC of 0.968 for the sparse model, with an estimated optimism of 0.007. For comparison, XGBoost showed an apparent AUC of exactly 1.000 with corrected AUC 0.984 (optimism 0.016), reflecting complete in-sample memorisation. The small optimism of the sparse model is consistent with its low effective dimensionality, but its magnitude warrants caution: in this small-sample, near-separable regime an apparent AUC of 0.975 leaves little headroom for the bootstrap to detect optimism, so an estimate of 0.007 is plausibly an underestimate of true out-of-sample shrinkage rather than evidence of negligible overfit. The corrected figure should therefore be read alongside, not in place of, the cross-validated and temporal estimates.

### 3.6 Overfitting assessment and the parsimonious two-term model

Because the events-per-candidate ratio at the screening stage is low (68 events over 71 library terms ≈ 0.96), we quantified overfitting at the *fitting* stage along three complementary axes and pre-specified a parsimonious model restricted to the two bootstrap-stable interactions. First, events-per-estimated-parameter at the fitting stage is 9.7 for the seven-term model and 34 for the two-term model, the latter comfortably within conventional targets. Second, the van Houwelingen–Le Cessie heuristic shrinkage factor 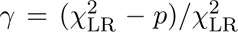 was 0.970 for the seven-term model and 0.991 for the two-term model, both above the 0.9 threshold below which Riley et al. flag material overfitting [1]. Third, the Riley (2019) minimum-sample-size criteria for a model of this dimensionality and apparent performance require *n* ≥ 64 (seven-term) and *n* ≥ 20 (two-term); the available *n* = 259 exceeds both, so the sample-size tension is confined to the 71-term *screening* library and does not apply to the deployed low-dimensional model. We note explicitly that these fitting-stage diagnostics do not themselves capture selection-induced optimism, which is addressed separately by the selection-inclusive bootstrap of §3.2 and the Harrell–Steyerberg correction above.

The pre-specified two-term model (intercept, dse×dshb, tvol×star) reproduced the discrimination of the full seven-term model: apparent C-statistic 0.970, 5 × 10 cross-validated C-statistic 0.968 per-fold and 0.964 pooled, with a Harrell–Steyerberg bootstrap optimism of only 0.002 (optimism-corrected 0.968). Its cross-validated calibration was at least as good as the seven-term model’s: calibration-in-the-large 26.2% predicted versus 26.3% observed, cross-validated calibration slope 0.926, and a bootstrap-validated calibration slope (uniform shrinkage factor) of 0.93 (interquartile range 0.80–1.08). Finally, an unpenalised maximum-likelihood fit of the seven-term model showed no quasi-separation (maximum standardised coefficient 4.24, well below the magnitude that signals separation, with finite converged estimates), indicating that the near-unity upper confidence bounds reflect a genuinely high signal-to-noise ratio in this measurement-consistent cohort rather than a numerical artefact. The two-term model therefore serves as a robustness analysis confirming that the deployed seven-term equation (Eq. (2)) is not an artefact of its non-stable terms. We nonetheless deploy the seven-term equation rather than the two-term model, because three of the five verified properties (P3–P5)—in particular the direction-of-effect of thyromental distance (P3)—concern terms present only in the fuller model; the two stable interactions are treated as its load-bearing core and the remaining five terms as transparent secondary structure.

### 3.7 Calibration

Calibration was assessed on the prevalence-true probability scale, with mean predicted probability against observed prevalence as the primary metric, supplemented by the calibration slope, the Brier score and the reliability diagram (Figure 3, Table 4); we deliberately avoid headlining the single-number Cox calibration-in-the-<u>large in</u>tercept, because for a highly discriminative model the Jensen gap between logit(*y̅*) and 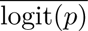 inflates it into an unreliable miscalibration signal (it is reported once in the Supplement with this caveat). On this scale calibration was good. In pooled 5×10 cross-validation the mean predicted probability was 26.2% against an observed prevalence of 26.3% (a near-perfect agreement in calibration-in-the-large), the calibration slope was 0.90, and the Brier score was 0.033 — far below the value of approximately 0.19 for a null model predicting the marginal prevalence. On the 80/20 test set the mean predicted probability was 26.5% against 26.9% observed, with Brier 0.028. The test-set calibration slope was 0.71; this is a substantial departure from the ideal value of 1.0, indicating that predicted risks on the small test partition (14 events) are over-dispersed and would benefit from shrinkage before deployment, and it should not be conflated with the well-behaved cross-validated slope of 0.90. Under the temporal hold-out (first enrolment half training, second testing; *n* = 129, 31 events) the over-extremity intensified rather than attenuating: calibration slope was 0.625 and Brier 0.052, indicating that predictions are increasingly mis-scaled under prospective shift even though the mean predicted risk continues to track prevalence approximately. This is the calibration signal most relevant for prospective deployment and reinforces that shrinkage recalibration on local data is required prior to clinical use. Bin-level observed-versus-expected agreement across risk strata is shown in the reliability diagram (Figure 3): the points track the identity line in the cross-validated panel, with the wider scatter on the test panel reflecting the small event count rather than directional bias.

**Figure 3:**
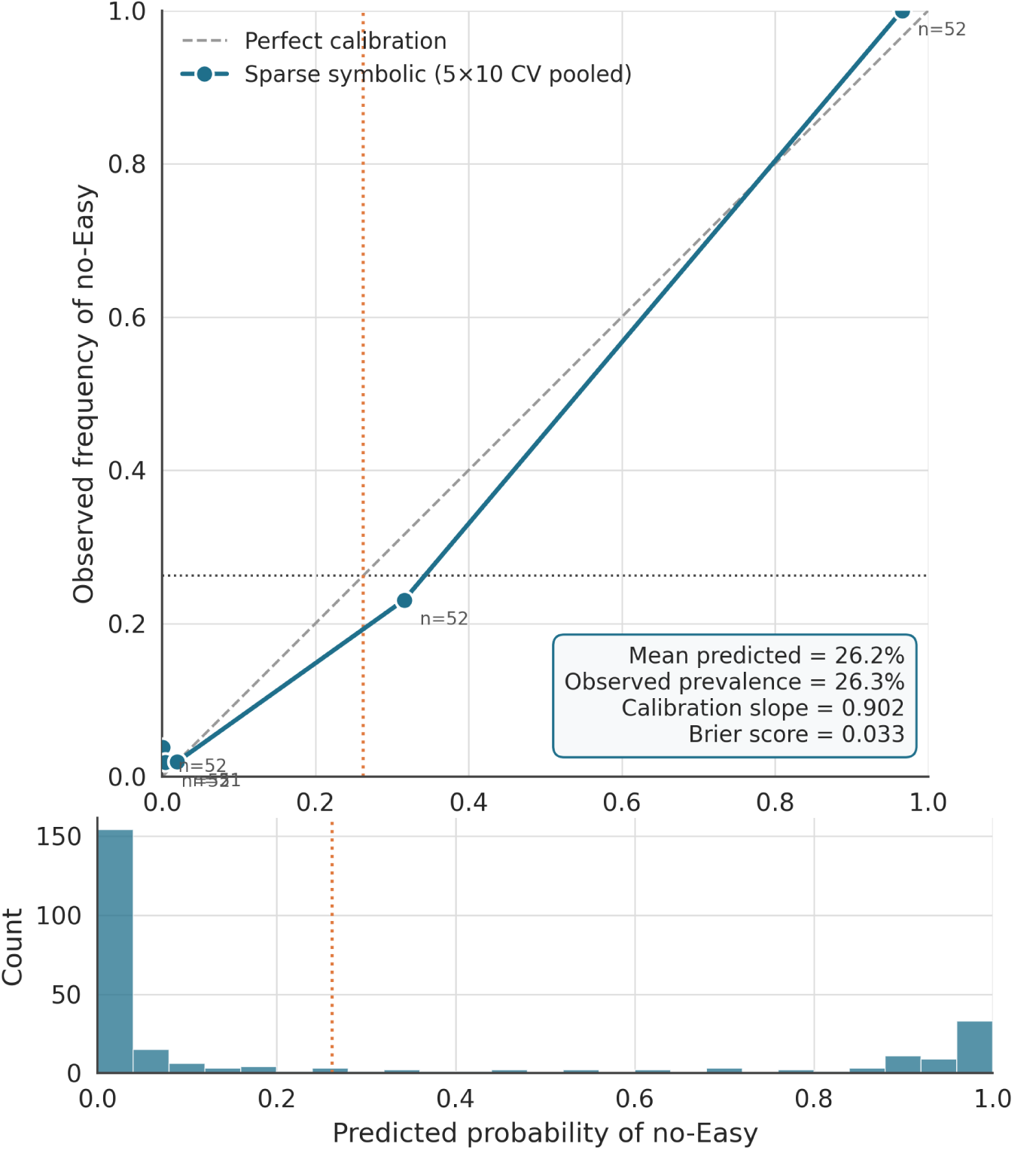
Reliability diagram of the Option-A sparse symbolic model on 5×10 cross-validated pooled predictions (five quantile bins), with the predicted-probability histogram below; the calibration curve tracks the identity line with mean predicted risk 26.2% essentially equal to the observed prevalence of 26.3%, calibration slope 0.904 and Brier score 0.033 (cross-validation), consistent with adequate calibration-in-the-large with mild over-extremity after the prevalence-true intercept refit; the more pronounced over-extremity on the 80/20 test (slope 0.71) and temporal hold-out (slope 0.625) is reported in the main text.

**Table 4:** Calibration and clinical utility of the sparse symbolic model. Panel A reports calibration evidence framed as calibration-in-the-large (mean predicted risk vs. observed prevalence), calibration slope, and Brier score. Panel B reports reclassification versus the clinical baseline (80/20 test) as categorical NRI (threshold 0.3), continuous NRI, and IDI. The categorical NRI is identical for both models (+0.444): a genuine coincidence at the single 0.3 boundary, disambiguated by the distinct continuous-NRI and IDI values.

| **Panel A. Calibration** |  |  |  |  |
| --- | --- | --- | --- | --- |
| Setting | Mean predicted (%) | Observed prevalence (%) | Slope | Brier |
| $5 \times 10$ CV (pooled) | 26.2 | 26.3 | 0.904 | 0.033 |
| 80/20 test | 26.5 | 26.9 | 0.714 | 0.028 |
| **Panel B. Reclassification vs. clinical baseline** |  |  |  |  |
| Model | Categorical NRI (0.3) |  | Continuous NRI | IDI |
| Sparse symbolic | +0.444 |  | +1.80 | +0.533 |
| LR (9 POCUS) | +0.444 |  | +1.86 | +0.536 |

NRI = net reclassification improvement; IDI = integrated discrimination improvement. Categorical NRI is evaluated at the single 0.3 risk threshold; the two models share the identical value +0.444 because they reclassify exactly the same test patients across that one boundary — a genuine coincidence, not a transcription error, as confirmed by the distinct continuous-NRI and IDI values that use the full predicted-probability distribution. Calibration slope of 1.0 is ideal; values $< 1$ indicate slightly over-extreme predictions. Brier score: lower is better (null model at 26% prevalence $\approx 0.19$ ). A reliability diagram is provided in the Supplementary Material; the Cox calibration-in-the-large intercept is reported there with the Jensen-gap caveat. Decision-curve analysis showed positive net benefit over the $\sim 10$ –50% threshold range, exceeding treat-all and treat-none strategies.

**Figure 4:**
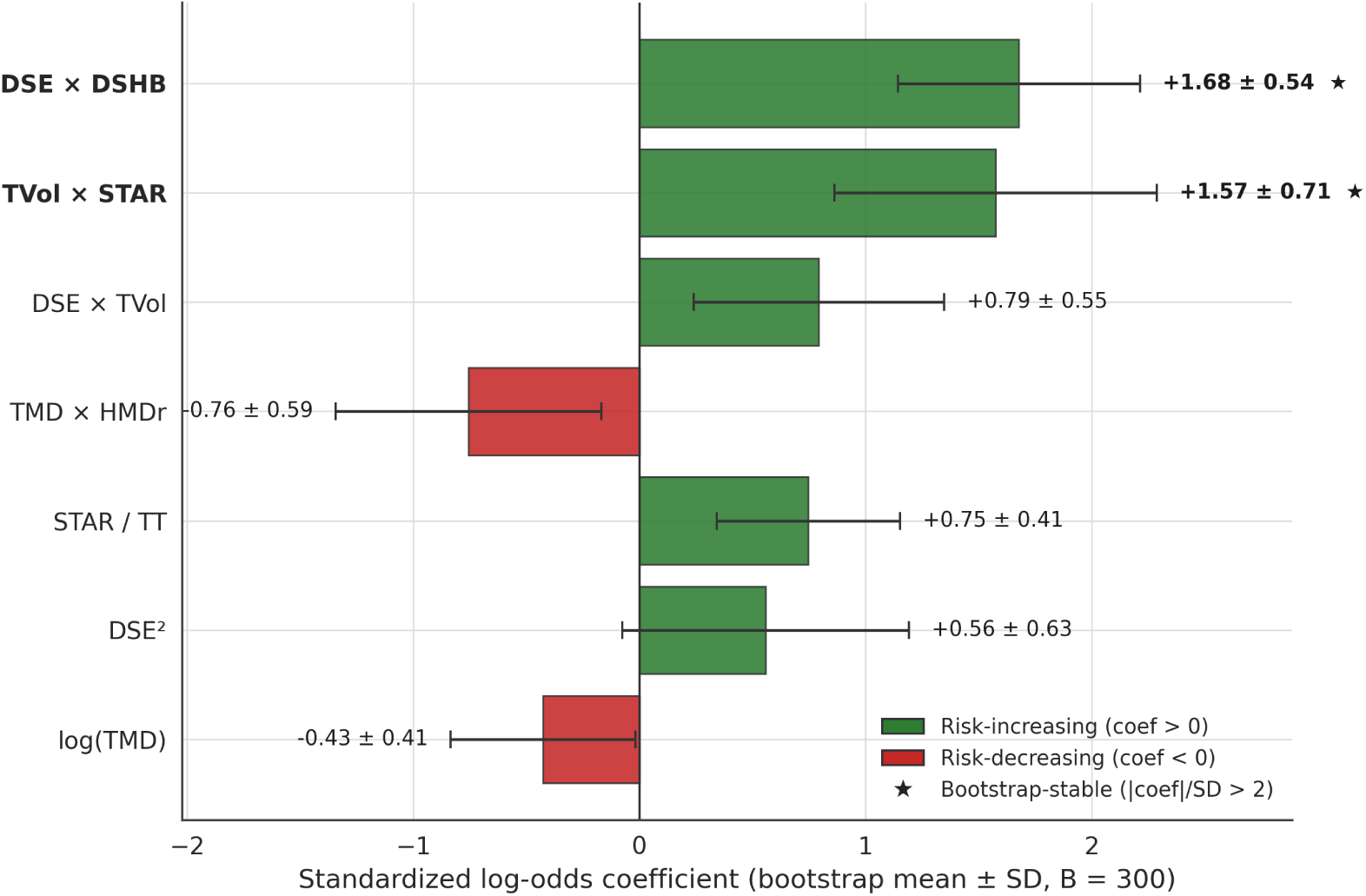
Standardized log-odds coefficients (bootstrap mean ± SD, B=300) of the seven retained sparse symbolic terms, ordered by magnitude and coloured by direction of effect (green risk-increasing, red risk-decreasing); the two bootstrap-stable interactions DSE × DSHB (+1.68±0.54; pre-laryngeal soft-tissue depth coupling, anterior neck) and TVol × STAR (+1.57±0.71; lingual-mass coupling), both with |coef|*/*SD *>* 2, are starred as the dominant, reproducible drivers of difficult airway risk.

To establish whether the temporal miscalibration reflects a loss of model usefulness or a correctable re-scaling, we applied standard logistic recalibration on the prospective half (*n* = 129, 31 events, prevalence 24.0%). Discrimination was unaffected by recalibration (AUC 0.941, necessarily unchanged because recalibration is monotone), so the ordering of patients transfers intact across time. An *out-of-sample* recalibration—5 × 10 cross-recalibration confined to the second half, so the intercept-and-slope map is never fitted and evaluated on the same patients—restored the calibration slope from 0.625 to 0.92 and the calibration-in-the-large intercept from +1.55 to approximately zero, realigning the mean predicted risk with observed prevalence (24.1% vs 24.0%). The temporal drift is therefore an affine re-scaling of an otherwise well-ordered predictor, correctable by the routine intercept-and-slope recalibration that TRIPOD+AI recommends at the point of deployment, rather than a failure of the model to transport across time. We nonetheless retain external, prospectively recalibrated validation as a prerequisite for clinical use.

### 3.8 Reclassification and decision curve analysis

Against the clinical baseline restricted to standard bedside predictors, the sparse symbolic model improved reclassification on the 80/20 test set across all three indices (Table 4): categorical Net Reclassification Index (NRI; threshold 0.30) +0.444, continuous NRI +1.80, and Integrated Discrimination Improvement (IDI) +0.533. The benchmark nine-predictor logistic regression yielded categorical NRI +0.444, continuous NRI +1.86, and IDI +0.536. The categorical NRI is numerically identical between the two models (+0.444 each): this is a genuine coincidence, not a transcription error, because at the single 0.30 boundary both models happen to reclassify exactly the same test patients; the distinct continuous-NRI and IDI values, which use the full predicted-probability distribution rather than one threshold, separate the two models and confirm the coincidence is threshold-specific. These are bare point estimates computed on a small test partition (52 patients, 14 events), and they should be read with corresponding caution: the category-based NRI is known to be unstable and upward-biased in small-event settings, and with only 14 events its sampling variability is large. We therefore report the NRI and IDI as descriptive, threshold-specific summaries rather than as confirmatory effect sizes, and weight the decision-curve evidence accordingly. Decision curve analysis (Figure 5) showed positive net benefit of the sparse model across the threshold range 10–50%, exceeding the clinical baseline and the “treat-all” / “treat-none” reference strategies throughout the clinically relevant range, with the advantage largest near the prevalence-aligned threshold (25–30%) at which a clinician would prepare advanced airway equipment.

**Figure 5:**
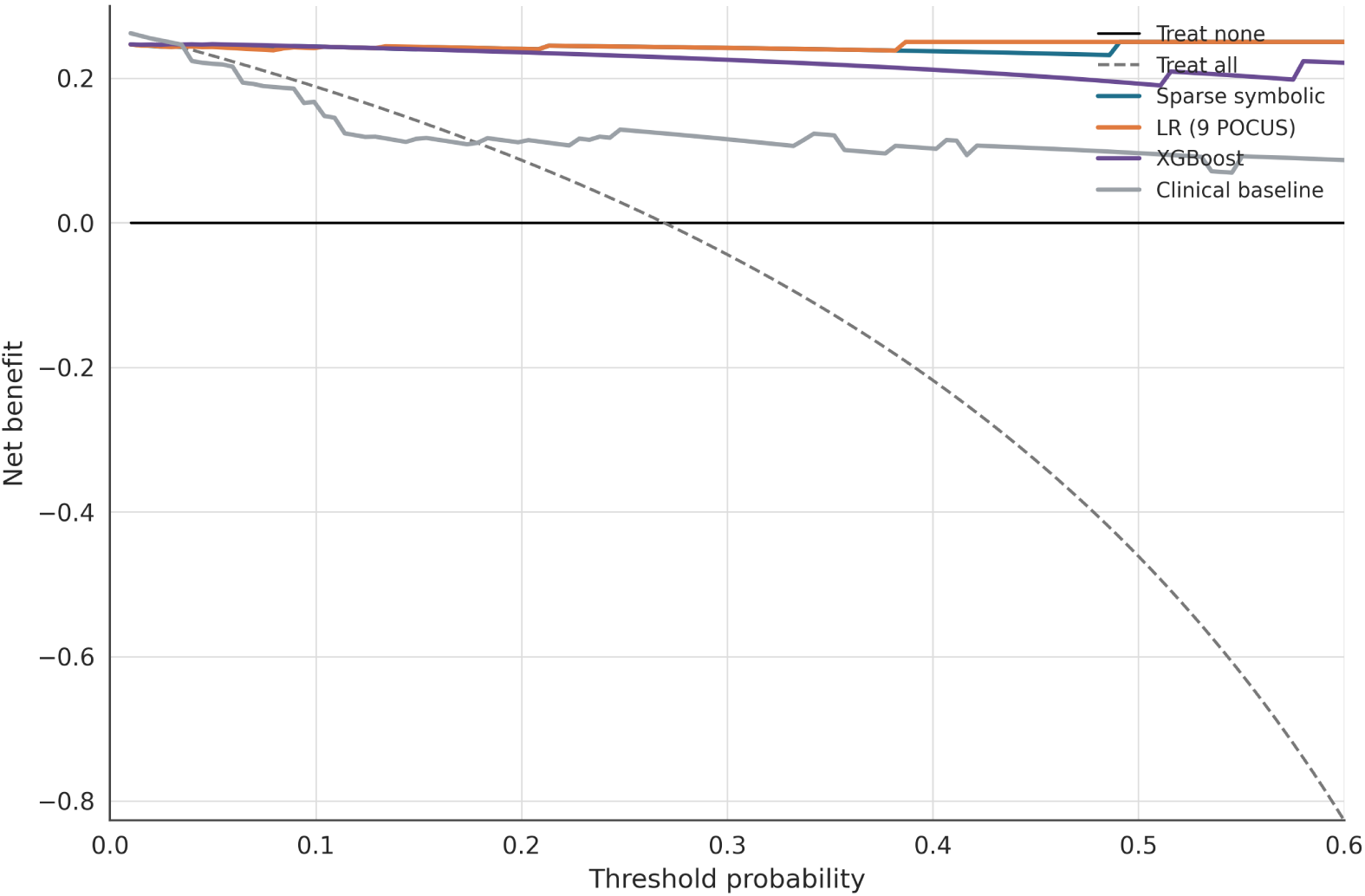
Decision-curve analysis on the 80/20 test set showing that the sparse symbolic, LR and XGBoost models yield higher net benefit than the treat-all and treat-none strategies and than the clinical baseline across the clinically relevant threshold range (∼10–50%), supporting net clinical utility of POCUS-based risk stratification.

### 3.9 Formal verification

The closed-form sparse symbolic predictor was transcribed into Lean 4 as a total function over a structure of nine real feature fields (with positivity supplied as an explicit hypothesis where required), and five theorems were machine-checked against the Mathlib library. The build completed successfully with zero sorry placeholders, certifying that the proofs are total: the stated properties hold for every admissible input vector rather than only on the sampled data. We stress that these are structural and mathematical properties of a *fixed* scoring function (its sign, domain and monotonicities); they are not claims of a clinical or causal effect, and they do not certify the empirical bootstrap-stability findings, the upstream reliability of the ultrasound measurements, or the calibration reported above.

The five verified theorems are: (P1) *sigmoid monotonicity* — the predicted probability is a strictly increasing function of the linear score, so the calibrated output respects the ordering induced by the symbolic decision function; (P2) *domain well-posedness* — the predictor is defined, finite and lies in (0, 1) for every input with strictly positive feature values, with no division-by-zero or logarithm-of-non-positive pathology on the admissible domain; (P3) *TMD direction-of-effect* — the predicted probability is strictly antitone in the sonographic thyromental distance over its positive domain, a statement holding across the entire admissible domain—the score decreases in TMD with the remaining features held fixed (a property of the scoring function, not an assertion that the variable is clinically protective); and (P4–P5) *sign and monotonicity of the bootstrap-stable interactions* — the score is strictly monotone in the DSE×DSHB and TVOL×STAR terms in the directions implied by their fixed positive coefficients. Together these theorems certify that the deployed formula cannot silently violate its specified qualitative structure under refactoring, retraining on the same library, or numerical re-implementation in a downstream language — a guarantee about the function that is complementary to, not a substitute for, the statistical validation above.

## 4 Discussion

### 4.1 Principal findings

To our knowledge, and scoped to a deployable clinical risk score, this work is the first to combine sparse symbolic regression with machine-checked formal verification: we are not aware of a prior clinical artificial intelligence (AI) predictor whose structural properties have been certified by a proof assistant rather than only empirically validated. We make this priority claim solely to the best of our knowledge, since the formal-methods and clinical-AI literatures are too large to have been searched exhaustively. Combining STLSQ [7], bootstrap stability selection, and theorem proving in Lean 4 with Mathlib [11, 12], we obtained an interpretable closed-form predictor of perioperative airway difficulty with high discrimination (5 × 10 cross-validated AUC 0.966 [0.926–1.000]; temporal hold-out 0.941; internal device hold-outs 0.947 and 0.972), adequate calibration-in-the-large with mild over-extremity, and machine-checked correctness over its full input domain. The optimism-corrected AUC of 0.968 (apparent 0.975, Harrell–Steyerberg optimism 0.007; *B* = 200) indicates that the seven-term predictor is not an artefact of an aggressive search. Only two retained terms met the pre-specified |*c*|*/σ_c_ >* 2 stability criterion, the interactions dse×dshb (3.13) and tvol×star (2.21); the other five (star*/*tt, dse×tvol, tmd×hmdr, dse^2^, log tmd) are retained as transparent secondary structure, not load-bearing components, as discussed under Limitations. The two stable interactions recover interpretable pre-laryngeal soft-tissue constructs (the skin-to-epiglottis × skin-to-hyoid distance coupling, which characterises the anterior soft-tissue column the blade must displace, and the tongue-volume × sagittal-tongue-area product, which characterises the lingual mass), consistent with the formally verified direction of effect.

### 4.2 Methodological contribution beyond explainability

Our contribution extends the frame of Rudin [4]: in high-stakes decisions, post-hoc explanations of opaque models are an inferior substitute for inherently interpretable ones. Tools such as SHAP [5] and LIME [6] explain a single prediction *a posteriori*; the five Lean 4 theorems we prove are instead quantified over *all* admissible inputs (domain admissibility, strict monotonicity in clinically directed features, and the sign of the dominant interaction), mechanically verified by a kernel that is itself the object of decades of metatheoretic scrutiny. Where testing rules out specific failures, verification rules out classes of them. This is not ornamental: a plausible re-implementation error—a flipped sign on the thyromental-distance coefficient, which would reverse the clinically established direction of effect—causes theorem P3 to fail to compile (Supplementary Material), so such an error cannot ship silently in the deployed artefact, whereas it could pass unit tests that do not probe monotonicity across the full input domain. We stress the exact content of theorem P3: it establishes that the predictor is strictly monotonically decreasing in thyromental distance (TMD) *by construction of the proof*, a property of the fitted function under its retained coefficients. It does not confirm a causal or protective effect of TMD in patients, which would require interventional or external evidence; the guarantee concerns the function’s monotonicity, not a patient-level effect. This is, in our reading, the methodological core of the manuscript; the airway use case is its first instantiation.

### 4.3 Methodological rigor and the auditable warrant of model claims: why equivalence in AUC is not equivalence in rigor

The numerical proximity of the three families of predictors in this cohort masks a substantive difference in methodological rigor that is in fact the contribution of this work. We compare them along four operational axes: (i) whether the functional form is pre-specified or recovered from data; (ii) whether the fitted model is identifiable as a closed-form expression that a third party can re-derive; (iii) whether the selection of structure is reproducible under perturbation of the training set; and (iv) whether behavioural properties of the predictor are verifiable over the feasibility region or only estimable on samples.

Penalised logistic regression fixes the functional form—linear-additive in the nine features—before the data are seen. Estimation then proceeds within that pre-specified structure, so the data are not used to question whether linearity is appropriate, only to identify coefficients within it. This yields an identifiable closed form (axis ii) but at the price of an untested specification (axis i): there is no axis-i evidence that the linear-additive form is the correct one for this prediction problem [14]. Gradient-boosted trees occupy the opposite end. The ensemble is selected from a high-capacity space of tree combinations that is not identifiable as a closed-form expression (axis ii): two equally performing ensembles need share neither variables nor split points, so the fitted model cannot be audited as a single mathematical object [4]. Selection stability is correspondingly weak (axis iii): the same boosting procedure on bootstrap resamples yields different feature-importance orderings without a quantitative resampling guarantee. Behavioural properties of the function—monotonicity, bounded output, sign of an effect—cannot be verified over the feasibility region (axis iv); they can only be probed empirically on the available sample, and post-hoc explanation tools (SHAP, LIME) produce local approximations whose stability is itself a known concern.

Sparse symbolic regression with formal verification addresses all four axes simultaneously. The functional form is *recovered* from a structured candidate space (originals, squares, pairwise and triple products, ratios, logarithms; axis i): which terms enter the model is an outcome of inference. The fitted model is *identifiable* as a seven-term closed-form logistic score whose intercept, coefficients and standardisation constants are fully disclosed (Eqs. (2)–(3), axis ii). The *stability* of structure selection is quantified by bootstrap resampling (*B* = 300): the two retained interactions that meet the pre-specified |*c*|*/σ_c_ >* 2 criterion (dse×dshb, ratio 3.13; tvol×star, ratio 2.21) constitute a testable claim about which mechanistic structures the data license—a claim that can be confronted with, and rejected by, independent multicentre data if the same interactions and signs do not resurface (axis iii) [10]. Finally, behavioural properties of the fitted predictor are *verified* over its feasibility region by five machine-checked Lean 4 theorems (P1–P5), so monotonicity in TMD, positivity of the dominant interaction coefficient and output domain (0, 1) are guaranteed for every admissible input rather than only observed on the cohort sample (axis iv).

The practical consequence for clinical deployment is not aesthetic. A predictor whose behavioural properties are only empirically observed transfers the verification burden to the clinician on every new patient, since out-of-sample inputs carry no guarantees. A predictor whose behavioural properties are mathematically certified over the feasibility region allows the clinician to rely on those properties without rechecking them case by case, and allows a regulator or institutional reviewer to audit them once and for all. Equivalence in AUC across the three families (Tables 3–4) should therefore not be read as evidence that the choice is indifferent. Discrimination is bounded above by the information content of the features; what differs across the three families is the auditable warrant the model can offer for the clinical act [2, 4]. In safety-critical settings the recover-identify-stabilise-verify pipeline is, on our reading, the methodologically warranted default whenever it is attainable at no discriminative cost, as is the case here.

### 4.4 Why a single-operator cohort is a strength for the methodological claim

A single-operator, single-centre design limits external validity, which we accept for any deployment claim and address under Limitations. For the methodological claim, however, the single-operator setting is informative: inter-operator variability in point-of-care airway ultrasound is a dominant source of nuisance noise, and eliminating it by design isolates the intrinsic discriminating power of the pipeline under measurement-consistent conditions, analogous to a controlled laboratory experiment. We stress that this yields a best-case-measurement-consistency estimate, not a generalisable performance ceiling: fixing the operator removes inter-operator variance but bakes in one expert’s idiosyncratic skill and bias, which could inflate or deflate discrimination relative to multi-operator practice. Multicentre, multi-operator validation is therefore the natural and intended next step.

### 4.5 Comparison with XGBoost benchmark

In-cohort, the sparse symbolic predictor (CV AUC 0.966 [0.926–1.000]) was numerically comparable to a tuned XGBoost benchmark (CV AUC 0.955) and to penalised logistic regression (0.967), and exceeded the clinical baseline (0.809). The pattern was consistent across resampling protocols: XGBoost device hold-outs were 0.949 (McGrath→C-MAC) and 0.969 (C-MAC→McGrath) and its temporal hold-out 0.948, against sparse values of 0.947, 0.972 and 0.941 respectively. We refrain, however, from asserting statistical equivalence or superiority: no DeLong or paired-bootstrap test was performed, the bootstrap intervals are wide (sparse CV CI 0.926–1.000), and the apparent margin over the clinical baseline was not subjected to an inferential paired comparison. The differential we do claim is epistemic rather than discriminatory: the sparse model offers (i) an explicit closed-form formula amenable to direct clinical inspection; (ii) machine-verifiable structural properties; and (iii) parsimony, with seven retained terms versus 300 trees in the boosted ensemble. The bias–variance trade-off that justifies tree ensembles cannot, by itself, substitute for the absence of formal guarantees in safety-critical settings.

### 4.6 Calibration and clinical utility

Calibration was assessed under a prevalence-true (intercept-anchored) fit. Calibration-in-the-large was adequate: mean predicted risk matched observed prevalence in cross-validation (26.2% vs 26.3%) and on the 80/20 test split (26.5% vs 26.9%). Because the intercept was anchored to prevalence, this near-match is partly a consequence of the fitting choice and is not, by itself, independent evidence of fine-grained calibration. The calibration slope was acceptable in cross-validation (0.904) but fell to 0.714 on the smaller test split, indicating predictions that are somewhat too extreme out of sample; the Brier score remained low (0.033 cross-validation, 0.028 test) against a ∼0.19 no-information reference at the observed prevalence. We therefore characterise calibration as adequate calibration-in-the-large with mild over-extremity (slope *<* 1), as visualised in the reliability diagram (Figure 3), rather than as strong or near-perfect [1, 2]. Against the clinical baseline, reclassification favoured the symbolic model (categorical NRI +0.444, continuous NRI +1.80, IDI +0.533); we report these with the caveat that NRI and IDI are known to be unstable in modest samples and read them as supportive rather than confirmatory. Decision-curve analysis showed positive net benefit across thresholds of approximately 10–50%, exceeding treat-all and treat-none references over the clinically relevant range.

### 4.7 Generalisation beyond airway difficulty

The pipeline (STLSQ with bootstrap stability selection, Lean verification of structural properties, and prevalence-true recalibration) applies in principle to any clinical prediction problem with continuous features admitting a meaningful feasibility region, a population manifold smooth enough for a sparse polynomial-plus-ratio library to be expressive, and plausible parsimonious mechanistic structure. Candidate domains include intraoperative haemodynamic risk, sepsis early-warning scores, and oncologic surrogate endpoints. Extensions to dynamics through the full SINDy and E-SINDy formalism [8] and to hybrid universal differential equations [9] are the natural research trajectory.

### 4.8 Limitations

Several limitations qualify our conclusions. First, the cohort is single-centre and acquired by a single operator (MAFV); the temporal and device hold-outs are internal resampling splits of this same single-centre cohort, not external validation, and the “device-transfer” label denotes a McGrath/C-MAC partition (132/127) rather than an independent site. Multicentre external validation remains the next step; a companion analysis of the three-centre extension of this cohort (Oyarzún-Silva et al., in preparation) shows that ultrasound predictive signal does not transport across non-harmonised operators and localises the failure to measurement acquisition rather than to the model or the populations, so external validation must be preceded by prospective measurement harmonisation and operator credentialing. Because the same operator acquired the predictors and was not blinded to them when grading the outcome, incorporation bias cannot be excluded; we note, however, that the deployed score discriminated the most objective outcome component—escalation to a hyperangulated device (grade 2)—with a cross-validated C-statistic of 0.926 and ranked the three difficulty grades monotonically (Spearman *ρ* = 0.71, *p <* 10^−30^), which argues that the association reflects a documented procedural reality rather than the single rater’s subjective impression alone; blinded, multi-operator assessment would nonetheless be required to settle this definitively. Second, the sample size (*n* = 259 with 68 events) is modest, and with a 71-term candidate library the events-per-candidate-parameter ratio during selection is low; although bootstrap stability and optimism correction support the two-term core, the risk of overfitting during feature selection is real, which is precisely why only the two stability-selected terms are treated as load-bearing [1]. That five of the seven retained terms fail the |*c*|*/σ_c_ >* 2 threshold tempers, rather than supports, a strong parsimony-and-robustness narrative. Third, the Lean 4 verification certifies structural properties of the predictor as a mathematical object; it cannot certify the upstream reliability of the ultrasound measurements, which remains empirical. Fourth, although prespecified fairness analyses report discrimination and calibration by sex and BMI stratum (Supplementary S3), these subgroups were not separately powered—the obese stratum is small (*n* = 37, with an unstable calibration slope)—and device subgroups beyond the McGrath/C-MAC partition were not examined; the subgroup calibration differences (notably under-prediction in women) reinforce that local recalibration is required before use. Fifth, decision-threshold optimisation was deliberately out of scope; deployment would require local threshold tuning under the prevalence and utility structure of the receiving institution, as recommended by TRIPOD+AI [3]. The full set of retained coefficients, the operating definition of each of the nine ultrasound features, the statements and complete Lean 4 proofs of the five theorems, the analysis scripts, and runnable pseudocode of the STLSQ pipeline are provided in the manuscript, Supplementary Material and public code repository, so that the model equation and its verified properties can be inspected and reproduced independently; only the STLSQ screening hyperparameters remain under a research collaboration agreement while the patent application is pending.

### 4.9 Conclusion

The combination of sparse symbolic regression with formal verification in Lean 4 produces a clinical risk score whose specification is recovered from data rather than pre-assumed, whose closed form is fully identifiable and disclosed, whose structure-selection stability is quantified by bootstrap resampling, and whose behavioural properties are mathematically certified over its feasibility region rather than only sampled empirically—at no demonstrable discriminative cost relative to penalised logistic regression and gradient-boosted-tree alternatives. Equivalence in AUC across these three families is, on our reading, precisely the condition under which the methodological-rigor differences among them become the decisive criterion rather than an ancillary one: when discrimination is bounded above by the information content of the features, what distinguishes competing predictors is the strength of the auditable warrant they can offer for the claims they make about the prediction problem. We propose this recover-identify-stabilise-verify pipeline as a natural complement to TRIPOD+AI-conformant reporting in safety-critical settings—perioperative airway management, intensive-care early warning, oncologic decision support—pending the multicentre, multi-operator external validation that these single-centre results motivate but cannot themselves provide.

## Supporting information

TRIPOD+AI 2024 reporting checklist

## Data Availability

The deployable equation, its standardisation constants and the five Lean 4 theorem statements are disclosed in full in the manuscript. The complete Lean 4 proof project and the analysis, validation and figure-generation scripts are publicly available at https://doi.org/10.5281/zenodo.22143665 under the PolyForm Noncommercial License 1.0.0; that repository includes a script that reproduces and verifies the deployed equation without the protected screening hyperparameters. The de-identified analysis dataset is under controlled access governed by the institutional data-access committee at Universidad de Navarra; requests are processed within 30 days for non-commercial research use. The two STLSQ screening hyperparameters remain available under a research collaboration agreement while the patent application covering the screening methodology is pending; they govern model discovery only.

## Supplementary Materials

The following supporting information is available: Algorithm S1, runnable pseudocode of the STLSQ + bootstrap stability-selection pipeline; the complete Lean 4 proof of Theorem P1 (sigmoid monotonicity) as a reproducibility demonstrator; intended use, target population and model-lock information; the TRIPOD+AI 2024 checklist; the reliability diagram and the Cox calibration-in-the-large intercept with its Jensen-gap caveat; and prespecified fairness analyses by sex and BMI stratum.

## List of abbreviations

AUC: area under the receiver operating characteristic curve
BMI: body mass index
CI: confidence interval
CITL: calibration-in-the-large
CV: cross-validation
DCA: decision-curve analysis
DSE: distance from skin to epiglottis
DSHB: distance from skin to hyoid bone
EPV: events per variable
HMDr: hyomental distance ratio
IDI: integrated discrimination improvement
IID: inter-incisor distance
LR: logistic regression
MMS: modified Mallampati score
NC: neck circumference
NRI: net reclassification improvement
POCUS: point-of-care ultrasound
SMD: sternomental distance
STAR: sagittal tongue area
STLSQ: sequentially thresholded least squares
TMD: thyromental distance
TRIPOD+AI: transparent reporting of a multivariable prediction model for individual prognosis or diagnosis (artificial-intelligence extension)
TT: tongue thickness
TVol: tongue volume
TW: tongue width
ULBT: upper-lip bite test
VCI: video classification of intubation
VL: videolaryngoscopy
XGBoost: extreme gradient boosting.

## Authors’ contributions

Conceptualization, R.O.-S. and M.Á.F.-V.; methodology, R.O.-S.; software, R.O.-S.; formal analysis, R.O.-S.; investigation, M.Á.F.-V. and N.D.L.-C.; data curation, M.Á.F.-V.; writing—original draft preparation, R.O.-S.; writing—review and editing, P.H.-H., M.Á.F.-V. and N.D.L.-C.; supervision, N.D.L.-C. and M.Á.F.-V.; funding acquisition, N.D.L.-C. All authors read and approved the final submitted version and agree to be accountable for their contributions and for the accuracy and integrity of the work.

## Funding

Financial support for the publication of this article was received from Fondos Organización Sanitaria Integrada (OSI) Bilbao-Basurto. The funders had no role in study design, data collection and analysis, the decision to publish, or preparation of the manuscript.

## Ethics approval and consent to participate

The study was conducted in accordance with the Declaration of Helsinki and Good Clinical Practice, and approved by the Research Ethics Committee of the University of Navarra on 22 December 2022 (protocol code 2022.193 mod1); it was registered at ClinicalTrials.gov (NCT06925009); a separate study protocol was not published. Written informed consent to participate was obtained from all patients prior to enrolment.

## Consent for publication

Not applicable.

## Patient and public involvement

No patients or members of the public were involved in the design, conduct, reporting, or dissemination plans of this research.

## Availability of data and materials

The deployable equation, standardisation constants, and the five Lean 4 theorem statements are disclosed in full in this manuscript. The complete Lean 4 proof project and the analysis, validation and figure-generation scripts are released in the public repository airway-sparse-lean (https://doi.org/10.5281/zenodo.22143665) under the PolyForm Noncommercial License 1.0.0. The de-identified analysis dataset is deposited at Zenodo under controlled access governed by the institutional data-access committee at Universidad de Navarra (DOI to be issued at acceptance); requests are processed within 30 days for non-commercial research use. Only the exact STLSQ screening hyperparameters (*λ*, *τ*) remain under a research collaboration agreement while the patent application covering the screening methodology is pending; they affect model discovery only: the repository includes verify_equation_public.py, which reconstructs the seven disclosed terms, refits the deployed equation and checks it against both the published coefficients and the Lean constants without using them, and the Lean proof project re-verifies every formal guarantee from the repository alone, requiring neither the hyperparameters nor the data. Requests should be directed to the corresponding author.

## Acknowledgements

Not applicable.

## Competing interests

A patent application covering the methods described in this manuscript is pending. R.O.-S. and M.Á.F.-V. are named on this pending patent application and declare this as a competing interest. The remaining authors declare that they have no competing interests.

## Relationship to prior work

This study re-analyses an extended version of the single-operator cohort first reported in the parent Airway Coach study [13] (accepted, *BMC Anesthesiology*). The two works are non-redundant: the parent develops a three-class videolaryngoscopy-strategy classifier using conventional tree-ensemble, support-vector and multinomial-regression algorithms, whereas the present work addresses a binary *Easy*/*no-Easy* outcome with sparse symbolic regression, incorporates additional ultrasound-derived parameters, contributes a disclosed closed-form equation with formal (Lean 4) verification, and adds a TRIPOD+AI-conformant overfitting, internal-validation and calibration assessment. The overlap and the differences are disclosed here and in the cover letter.

## Declaration of generative AI and AI-assisted technologies

During the preparation of this work the authors used a large language model (Claude, Anthropic) to assist with language editing, LaTeX formatting, and the implementation and cross-checking of analysis scripts. All analytical results were independently verified against the source data by the authors, who reviewed and edited the content as needed and take full responsibility for the content of the publication.

## Notes

### Competing Interest Statement

A patent application covering the methods described in this manuscript is pending. R. Oyarzun-Silva and M.A. Fernandez-Vaquero are named on this pending patent application and declare this as a competing interest. The remaining authors declare that they have no competing interests. No author or institution received payments or services from a third party for any aspect of the submitted work in the past 36 months.

### Clinical Trial

NCT06925009

### Author Declarations

Ethical approval was GRANTED by the Research Ethics Committee of the University of Navarra (Comite de Etica de la Investigacion, Universidad de Navarra), Pamplona, Spain, on 22 December 2022, protocol ID 2022.193 mod1. The study was registered at ClinicalTrials.gov (NCT06925009). Written informed consent to participate was obtained from all patients prior to enrolment, and all procedures complied with the Declaration of Helsinki and Good Clinical Practice. The cohort was recruited at the Clinica Universidad de Navarra, Pamplona, Spain; this manuscript reports data from that single centre only.

