## Supplementary Material (S1-S7) for "An interpretable, formally verified point-of-care ultrasound risk equation for difficult videolaryngoscopy: development and internal validation"

This Supplementary Material accompanies the main manuscript. It contains: **S1** runnable pseudocode of the STLSQ + bootstrap stability-selection pipeline; **S2** a worked Lean 4 proof of Theorem P1 (sigmoid monotonicity), reproduced here for readers who will not build the full proof project; **S3** prespecified fairness analyses by sex and body-mass index; **S4** the Cox calibration-in-the-large intercept with the Jensen-gap caveat; **S5** a TRIPOD+AI 2024 reporting map; **S6** the intended use, target population and model-lock record; and **S7** the contents of the public code and proof repository.

---

#### S1. Algorithm: STLSQ with bootstrap stability selection

The procedure recovers a sparse symbolic logistic predictor from a candidate library and quantifies the stability of structure selection by resampling. It is given here at a level sufficient for independent re-implementation. The random seeds are included in the released code (S7); the exact sparsity threshold $\tau$ and regularisation strength $\lambda$ are available under a research collaboration agreement while the patent application covering the screening methodology is pending. These two values govern model *discovery* only, and the repository is built so that this restriction costs a reader nothing that matters. Scripts that re-run the screening stage require them and say so on exit; the script `verify_equation_public.py` does not: it rebuilds the seven *disclosed* terms, refits the deployed equation with the disclosed estimator settings, and checks the result against both the published values and the constants hard-coded in the Lean project. On the development cohort it reports 30/30 comparisons passing. The Lean proof project needs neither the hyperparameters nor the data: `lake build` re-verifies every formal guarantee from the repository alone.

```
INPUT : feature matrix X (n x 9), binary outcome y (n),
        threshold tau, L1 strength lambda, max iterations K,
        bootstrap count B, stability cutoff r = 2
------------------------------------------------------------------
LIBRARY construction Theta(X)  -> 71 columns
   winsorise each raw feature at 1st/99th percentiles
   columns = [ x_j ; x_j^2 ; x_i*x_j (i<j) ;
               8 pre-specified ratios x_a/x_b (denominator floored) ;
               log(x_j) ]           # ratios/logs on raw scale
   standardise every column to zero mean, unit variance (store mu, s)

PROCEDURE STLSQ(Theta, y, weights w):
   active <- all columns
   repeat up to K times:
       fit penalised logistic regression on Theta[:,active]
```

```
            (L1, strength lambda) with sample weights w
       drop columns with |coef| < tau from active
       if active unchanged: break
   refit UNWEIGHTED L2 logistic on Theta[:,active]   # Option A:
   return support 'active', coefficients, intercept   # prevalence-true

APPARENT model:
   w <- sqrt-balanced class weights
   (support*, coef*, b*) <- STLSQ(Theta, y, w)        # 7 terms

BOOTSTRAP stability selection (b = 1..B):
   draw stratified bootstrap resample (Theta_b, y_b, w_b)
   (support_b, coef_b, .) <- STLSQ(Theta_b, y_b, w_b)
   accumulate coefficient of each library term across resamples
for each retained term j:
   stable_j  <-  |mean(coef_b[j])| / SD(coef_b[j]) > r
------------------------------------------------------------------
OUTPUT: deployed 7-term equation (support*, coef*, b*, mu, s);
        bootstrap mean +/- SD and stability flag per term.
```

Internal validation re-executes the *entire* procedure (library construction and STLSQ selection) inside every cross-validation fold and every Harrell–Steyerberg bootstrap resample, so reported optimism reflects selection as well as estimation.

---

#### S2. Lean 4 proof of Theorem P1 (sigmoid monotonicity)

Theorem P1 states that the predicted probability is a strictly increasing function of the linear score $\eta$, i.e. the logistic link $\sigma(\eta) = (1 + e^{-\eta})^{-1}$ is strictly monotone. The script below is the exact proof from the verified project (the real type $\mathbb{R}$ is written `Real` for typesetting). The complete project—P1 through P5 over the nine-feature structure—compiles under Lean 4.30.0 with Mathlib and **zero `sorry` placeholders** (`lake build` succeeds).

```
import Mathlib.Analysis.SpecialFunctions.Exp

/-- Standard logistic sigmoid (the type Real is Lean's notation for the reals). -/
noncomputable def sigmoid (t : Real) : Real := 1 / (1 + Real.exp (-t))

/-- P1: the sigmoid is strictly increasing in its argument. -/
theorem sigmoid_strictMono : StrictMono sigmoid := by
  intro a b hab
  unfold sigmoid
  have hexpa : 0 < Real.exp (-a) := Real.exp_pos _
  have hexpb : 0 < Real.exp (-b) := Real.exp_pos _
  have hdb   : 0 < 1 + Real.exp (-b) := by linarith
  have hexp_lt : Real.exp (-b) < Real.exp (-a) :=
    Real.exp_lt_exp.mpr (by linarith)
  exact one_div_lt_one_div_of_lt hdb (by linarith)
```

The remaining theorems are proved in the same project: `predict_in_unit_interval` (P2, codomain $(0, 1)$); `predict_strictAnti_in_tmd` (P3, strict decrease in thyromental distance on the positive domain with hyomental-distance ratio $> 0$, since TMD enters through both the TMD×HMDR and $\log$ TMD terms, each with a negative coefficient); and `dse_dshb_term_strictMono` / `tvol_star_term_strictMono` (P4–P5, strict monotone increase of the two bootstrap-stable interaction contributions, matching their positive coefficients). A reviewer-confidential deposit of the complete proof project is provided during peer review.

**Why this is not ornamental: verification catches a silent error.** Consider a plausible re-implementation mistake—a flipped sign on the thyromental-distance coefficient (correctly −0.672, mistyped +0.672), which would make the model assert that a *longer* thyromental distance raises difficulty, the opposite of the established clinical direction. Such a typo can pass unit tests that do not probe monotonicity across the full input domain, yet it violates theorem P3: the sign obligation cannot be discharged and the build fails.

```
def betaTmdHmdrBugged : Real := 0.672    -- BUG: should be -0.672
theorem p3_sign_guarantee : betaTmdHmdrBugged < 0 := by norm_num
-- error: unsolved goals   |- betaTmdHmdrBugged < 0
```

Compilation halts, so the deployed artefact cannot silently ship with a clinically reversed effect. This is the concrete sense in which the formal layer *constrains*, rather than merely explains, the model.

---

#### S3. Prespecified fairness analyses by sex and body-mass index

Subgroup discrimination and calibration of the deployed seven-term model were evaluated on the pooled $5 \times 10$ cross-validation out-of-fold predictions (Table 1). Discrimination was preserved across all subgroups (C-statistic 0.94–1.00). Calibration, however, varied: the model *under-predicted* risk in women (mean predicted 10.9% vs observed 13.7%) and in lean patients, and *over-predicted* in the obese stratum, which is small ($n = 37$, 18 events) and yields an unstable calibration slope. These subgroup patterns mirror the overall out-of-sample over-extremity reported in the main text and reinforce that subgroup-aware recalibration on local data is required before deployment; no subgroup showed a discrimination failure.

**Objective-outcome check (incorporation bias).** As a probe of the single-operator, unblinded design, we evaluated how well the deployed no-Easy score discriminates the most *objective* outcome component—escalation to a hyperangulated device (grade 2), a documented procedural event. On pooled $5 \times 10$ cross-validation the score reached a C-statistic of 0.926 for grade 2 versus the rest, and predicted risk increased monotonically across grades (medians 0.7%, 93.4%, 99.3% for G0/G1/G2; Spearman $\rho = 0.71$, $p < 10^{-30}$). Discriminating a hard procedural event argues that the association reflects a documented reality rather than the rater's subjective impression alone, though it does not replace blinded, multi-operator validation.

Table 1: Subgroup discrimination and calibration (pooled $5 \times 10$ cross-validation, deployed seven-term model). Prevalence and observed are the subgroup event fractions; mean predicted is the mean cross-validated risk.

| Subgroup | $n$ | Events | C-statistic | Mean predicted (%) | Observed (%) | Calibration slope |
|---|---|---|---|---|---|---|
| Overall | 259 | 68 | 0.963 | 26.2 | 26.3 | 0.90 |
| Female | 117 | 16 | 0.939 | 10.9 | 13.7 | 0.84 |
| Male | 142 | 52 | 0.978 | 38.8 | 36.6 | 1.11 |
| BMI $< 25$ | 110 | 17 | 0.942 | 13.8 | 15.5 | 0.77 |
| BMI 25–30 | 112 | 33 | 0.962 | 29.1 | 29.5 | 0.97 |
| BMI $\geq 30$ | 37 | 18 | 0.997 | 54.6 | 48.6 | 2.86 |

The BMI $\geq 30$ subgroup ($n = 37$) is small; its near-unity C-statistic and inflated calibration slope are unstable and should be read as exploratory.

---

#### S4. Cox calibration-in-the-large and the Jensen gap

The main text reports calibration-in-the-large as mean predicted probability versus observed prevalence (26.2% vs 26.3% on pooled cross-validation). The Cox calibration-in-the-large intercept—the intercept $\alpha$ of a logistic regression of the outcome on the linear predictor entered as an offset—was $\alpha = +0.010$ on the pooled cross-validation, consistent with that agreement and indicating no mean miscalibration.

We caution against a naive alternative sometimes quoted as a "calibration-in-the-large" value, namely $\text{logit}(\bar{y}) - \overline{\text{logit}(p)}$. For a highly discriminative model whose predictions concentrate near 0 and 1, Jensen's inequality makes $\overline{\text{logit}(p)}$ strongly negative, inflating this difference to +1.69 here despite excellent mean calibration. This quantity is an artefact of the Jensen gap, not evidence of miscalibration, which is why the main text relies on the mean-predicted-versus-prevalence comparison and the offset intercept above.

---

#### S5. TRIPOD+AI 2024 reporting map

| TRIPOD+AI domain | Where addressed |
|---|---|
| Title / Abstract | Structured abstract; "development and internal validation" framing |
| Background / Objectives | Introduction |
| Source of data / Participants | Methods, *Cohort* (single centre; NCT06925009; data freeze) |
| Outcome | Methods, *Cohort* (Airway Coach grade; binary Easy/no-Easy; VCI framework) |
| Predictors | Methods, *Predictor variables* (nine POCUS features; definitions) |
| Sample size / EPV | Methods & Results, *Overfitting assessment* (EPP, shrinkage, Riley) |
| Missing data | Complete-case; one exclusion, no imputation |
| Statistical methods | Methods, *STLSQ*; *Validation pipeline*; S1 |
| Model specification | Results, deployed seven-term equation + standardisation table |
| Model performance | Discrimination, Calibration, Optimism, Reclassification, DCA |
| Model evaluation / fairness | S3 (subgroups by sex and BMI) |
| Interpretability / verification | Formal verification (Lean 4); S2 |
| Limitations | Discussion, *Limitations* |
| Data / code availability | Data Availability Statement; reviewer-confidential deposit |
| Funding / conflicts | Funding; Conflicts of Interest (patent disclosed) |

A fully itemised TRIPOD+AI 2024 checklist (item-by-item with page numbers) is provided as a separate completed form at submission.

---

#### S6. Intended use, target population, and model-lock

**Intended use.** The model is a pre-procedural risk-stratification aid: from nine point-of-care ultrasound measurements it estimates the probability that videolaryngoscopy will be *no-Easy* (require adjuncts or escalation to a hyperangulated blade), to support airway planning and resource preparation. It is a decision-support adjunct, not an autonomous or diagnostic device, and does not replace clinical judgement.

**Target population.** Adults ($\geq$ 18 years) scheduled for elective surgery requiring videolaryngoscopy-assisted orotracheal intubation, ASA physical status I–III, in whom standardised airway ultrasound is feasible. Excluded: emergency surgery, awake intubation, cervical immobilisation precluding ultrasound, and major airway pathology distorting baseline sonoanatomy. Use outside this population (e.g. emergency, obstetric, paediatric or critically ill patients) is not supported by the present evidence.

**Intended users and setting.** Anaesthesiologists and trained airway operators in the perioperative setting, using a comparable POCUS acquisition protocol; performance is conditional on measurement quality and was established with a single expert sonographer.

**Model-lock.** The deployed equation (intercept, seven standardised coefficients and the standardisation constants in the main-text table) is frozen; it is the full-cohort fit at the April 2026 data freeze. Version 1.0. Any re-estimation, library change, or recalibration constitutes a new model version and revokes the formal-verification guarantees, which are tied to the hard-coded coefficient signs. Local shrinkage recalibration before deployment is recommended (main text), and would be recorded as a recalibrated derivative of v1.0.

---

### S7. Public code and proof repository

The repository `airway-sparse-lean` accompanies this manuscript under the PolyForm Noncommercial License 1.0.0. It contains two parts.

**`lean/` — the complete Lean 4 proof project.** All five theorems compile against Mathlib with no `sorry` placeholders: `Predictor.lean` (the equation as a total function, with the intercept, the seven coefficients and the fourteen standardisation constants as hard-coded reals), `Properties.lean` (the five theorems), and `Examples.lean` (worked numerical instances). Re-verification is `lake exe cache get && lake build`: if it succeeds, the Lean kernel has re-checked every guarantee reported in the main text.

**`scripts/` — the analysis and validation pipeline.** Internal validation ($5 \times 10$ repeated cross-validation, temporal and device hold-outs), Platt and temporal recalibration, optimism correction, overfitting assessment, the Lasso and gradient-boosting benchmarks, the Youden operating points, the supplementary statistics of S3–S4, and the figure-generation code. Two scripts are verification rather than analysis: `verify_paper.py` refits the deployed equation from the raw data and compares the seven coefficients, means and standard deviations against the published values, and `33_verify_lean_vs_manuscript_vs_data.py` performs the three-way check *Lean constants == manuscript == recomputed-from-data*, which is the check that entitles a reader to trust that the equation proved in Lean is the equation reported here.

**Reproducing without the protected hyperparameters.** Scripts that re-run the screening stage read $\tau$ and $\lambda$ from a `screening_config.json` that is deliberately not distributed (S1) and exit with an explanatory message if it is absent. `verify_equation_public.py` is the exception and the one a reviewer needs: it rebuilds the seven disclosed terms from the raw measurements, refits the deployed equation with the disclosed estimator settings (unweighted $L_2$ refit, $C = 3.0$ — the soft class weights act during screening only), and checks intercept, all seven coefficients and all fourteen standardisation constants against both the published values and the constants hard-coded in `Predictor.lean`. On the development cohort it reports **30/30 comparisons passing** ($n = 259$, 68 events).
