## Supplementary material for "An interpretable, formally verified point-of-care ultrasound risk equation for difficult videolaryngoscopy: development and internal validation": TRIPOD+AI 2024 reporting checklist

This study is a prediction-model *development* study with internal validation (cross-validation plus temporal and device hold-outs); no external evaluation dataset was used, so items specific to model *evaluation* in a separate dataset are marked "Not applicable". Locations refer to manuscript sections; exact page/line numbers are to be inserted from the typeset proof. Items are from Collins GS, Moons KGM, Dhiman P, et al. *BMJ* 2024;385:e078378.

| Item | Checklist item (abbreviated) | Reported in |
|---|---|---|
| **TITLE & ABSTRACT** | | |
| 1 | Title: development/evaluation, target population, outcome | Title |
| 2 | Abstract (structured, TRIPOD+AI for Abstracts) | Abstract |
| **INTRODUCTION** | | |
| 3a | Background, healthcare context, rationale | Introduction (paras 1–3) |
| 3b | Target population, intended use, intended users | Introduction; Suppl. S6 |
| 3c | Known health inequalities between sociodemographic groups | No specific sociodemographic inequality in videolaryngoscopy difficulty was pre-specified; sex and BMI subgroups examined (Suppl. S3) |
| 4 | Objectives (development vs validation) | Introduction (objectives); Methods |
| **METHODS** | | |
| 5a | Source of data, rationale, representativeness | Methods, *Cohort* (prospective single-operator registry; extended extraction) |
| 5b | Dates of participant accrual | Methods, *Cohort* (from January 2023; April 2026 data freeze) |
| 6a | Setting, number/location of centres | Methods, *Cohort* (Hospital Universitario de Navarra, Madrid; single centre) |
| 6b | Eligibility criteria | Methods, *Cohort* (inclusion/exclusion) |
| 6c | Treatments received and handling | Methods, *Cohort* (videolaryngoscope device; adjunct/escalation use) |
| 7 | Data pre-processing and quality checking | Methods, *Sparse symbolic regression* (winsorisation, standardisation); *Cohort* (harmonised extraction) |
| 8a | Outcome definition, assessment, rationale | Methods, *Cohort* (Airway Coach grade; VCI framework; binary *Easy*/*no-Easy*) |
| 8b | Outcome assessor qualifications (subjective outcomes) | Methods, *Cohort* (single anaesthesiologist, >10 y airway-ultrasound experience) |
| 8c | Blinding of outcome assessment | Methods, *Cohort* (not blinded; incorporation bias flagged, mitigated by objective VCI procedural events) |
| 9a | Choice of initial predictors | Methods, *Predictor variables* (nine POCUS features; sonoanatomical rationale) |
| 9b | Predictor definitions, measurement | Methods, *Predictor variables* (definitions, units) |
| 9c | Predictor assessor qualifications | Methods, *Cohort*/*Predictor variables* (single operator) |
| 10 | Sample size and justification | Methods/Results, *Overfitting assessment* (events-per-parameter; Riley 2019 minimum sample size) |
| 11 | Missing data handling | Methods, *Cohort* (complete-case; one exclusion; no imputation) |
| 12a | Data partitioning, leakage avoidance | Methods, *Validation pipeline* ($5 \times 10$ CV, 80/20, temporal, device hold-outs; per-fold re-execution of all steps) |
| 12b | Predictor handling (transformation/standardisation) | Methods, *Sparse symbolic regression*; Results, standardisation table |
| 12c | Model type, building steps, internal validation | Methods, *Sparse symbolic regression* (STLSQ + bootstrap stability selection); *Validation pipeline*; *Formal verification* |
| 12d | Heterogeneity across clusters | Not applicable (single centre, single operator) |
| 12e | Performance measures and rationale | Methods, *Validation pipeline* (C-statistic, calibration, Brier, decision-curve analysis, NRI/IDI) |
| 12f | Model updating / recalibration methods | Results, *Calibration* (logistic recalibration on the temporal half) |
| 12g | (E) How predictions were calculated | Not applicable (development); deployable equation disclosed (Results, Eq. 1) |

| Item | Checklist item (abbreviated) | Reported in |
|---|---|---|
| 13 | Class imbalance methods and recalibration | Methods, *Sparse symbolic regression*/*Calibration* (square-root-balanced weights during selection; unweighted prevalence-true refit) |
| 14 | Fairness approaches and rationale | Suppl. S3 (subgroup discrimination/calibration by sex and BMI) |
| 15 | Model output (probability/classification, thresholds) | Methods, *Calibration* (probability); *Discrimination* (Youden threshold); Suppl. S6 |
| 16 | (E) Differences between development and evaluation data | Not applicable (development with internal validation only) |
| 17 | Ethical approval and consent | Declarations, *Ethics approval and consent to participate* |
| **OPEN SCIENCE** | | |
| 18a | Funding and role of funders | Declarations, *Funding* |
| 18b | Conflicts of interest | Declarations, *Competing interests* |
| 18c | Protocol availability | Declarations, *Ethics...* (no separate protocol published; prospectively registered) |
| 18d | Registration | Declarations, *Ethics...* (ClinicalTrials.gov NCT06925009) |
| 18e | Data sharing | Declarations, *Availability of data and materials* (controlled-access Zenodo deposit) |
| 18f | Code sharing | Declarations, *Availability of data and materials*; Methods, *Software* (versions); Suppl. S1 (runnable pseudocode); Suppl. S2 (Lean proof) |
| **PATIENT & PUBLIC INVOLVEMENT** | | |
| 19 | Patient and public involvement | Declarations, *Patient and public involvement* (none) |
| **RESULTS** | | |
| 20a | Participant flow (with/without outcome) | Results; Figure 1 (cohort flow) |
| 20b | Characteristics (incl. demographics, events, missing) | Results; Table 1 (baseline by grade) |
| 20c | (E) Comparison of development vs evaluation distributions | Not applicable (development) |
| 21 | Participants and events per analysis | Results (n and events per validation setting) |
| 22 | Full model specification (formula, code, access) | Results, *Final model specification* (Eq. 1 + standardisation table, fully disclosed); access terms in *Availability of data and materials* |
| 23a | Performance with CIs, key subgroups, plots | Results, *Discrimination*/*Calibration* tables (bootstrap 95% CI); Figures 2–3, 5; Suppl. S3 (subgroups) |
| 23b | Heterogeneity across clusters | Not applicable (single centre) |
| 24 | (E) Results of model updating | Results, *Calibration* (recalibrated temporal slope 0.92 out-of-sample) |
| **DISCUSSION** | | |
| 25 | Overall interpretation, fairness, prior studies | Discussion, *Principal findings*; *Methodological rigor* |
| 26 | Limitations (bias, sample size, overfitting, missing) | Discussion, *Limitations* |
| 27a | Handling poor/unavailable input data at deployment | Discussion, *Limitations*; Suppl. S6 (measurement quality) |
| 27b | User interaction and required expertise | Suppl. S6 (intended users; comparable POCUS protocol; expertise) |
| 27c | Next steps / generalisability | Discussion, *Limitations*/Conclusion (external, multi-operator validation) |

**Note.** Beyond TRIPOD+AI, the study additionally provides machine-checked formal verification of the deployed equation (Lean 4; Methods, *Formal verification*; Suppl. S2) and an explicit intended-use and model-lock record (Suppl. S6).
